# Distinct Sarcoma Microenvironments Predict Benefit from Addition of Pembrolizumab to Preoperative Radiotherapy and Surgery in SU2C-SARC032

**DOI:** 10.1101/2025.11.01.25339299

**Authors:** Stefano Testa, Jonathon E. Himes, Ajay Subramanian, Serey C.L. Nouth, Karla V. Ballman, Rachel S. Heise, Matthew Pierpoint, Neda Nemat-Gorgani, Timothy J. Sears, Michael S. Binkley, Anusha Kalbasi, David L. Corcoran, Angela M. Hong, Brian E. Brigman, Richard F. Riedel, Matt van de Rijn, Yvonne M. Mowery, Kent J. Weinhold, David G. Kirsch, Everett J. Moding

## Abstract

The addition of pembrolizumab to preoperative radiotherapy (RT) improved disease-free survival (DFS) for patients with stage III undifferentiated pleomorphic sarcoma (UPS) and dedifferentiated/pleomorphic liposarcoma (LPS) in the randomized SU2C-SARC032 trial. To precisely identify patients who benefit from pembrolizumab and RT, we performed comprehensive multi-omics profiling of pre- and post-treatment tumor and blood samples, including bulk RNA-seq, flow cytometry, and cytometry by time of flight. Additionally, we built a single-cell RNA-seq atlas spanning 65,786 cells from UPS and LPS to recover single-cell states in bulk tumor samples using digital cytometry. Two opposing tumor microenvironments (TMEs), immune-cold sarcoma ecotype 1 (SE1) and immune-hot sarcoma immune class E (SIC E), benefited from pembrolizumab. Pembrolizumab combined with RT caused an overall increase in activated CD8^+^ T cells, CD56^low^ NK cells, and T cell receptor diversity, while diminishing matrix-remodeling stromal cells and sarcoma cells. Our findings identify different mechanisms of response to pembrolizumab in localized, high-risk UPS/LPS and suggest that sarcoma TME signatures may identify patients most likely to benefit from adding pembrolizumab to preoperative RT.

## Introduction

Soft tissue sarcomas (STS) are a rare and heterogeneous group of mesenchymal malignancies^1^. Management for patients with high-risk, localized disease usually involves a combination of preoperative radiotherapy (RT) followed by surgery to optimize local control^2^. However, approximately half of patients with high-risk features, including large tumor size (> 5 cm) and high grade, ultimately develop metastatic disease^3^ with median overall survival of less than 2 years^4^. To reduce risk of metastasis, cytotoxic chemotherapy has been administered perioperatively to patients with high-risk localized STS with mixed results^5,6^. In the metastatic setting, immune checkpoint inhibition (ICI) targeting programmed cell death protein 1 (PD-1) and cytotoxic T-lymphocyte antigen-4 (CTLA-4) has shown promising activity in select STS histologies such as undifferentiated pleomorphic sarcoma (UPS) and pleomorphic or dedifferentiated liposarcoma (LPS)^7–9^. Furthermore, preclinical and clinical data have suggested that ICI can synergize with RT to improve local and distant anti-tumor responses^10,11^.

Recently, the SU2C-SARC032 phase 2 randomized trial investigated the benefit of adding perioperative ICI to preoperative RT and surgery for patients with Stage III (>5 cm and grade 2 or 3) UPS or LPS of the extremity or limb-girdle^12^. In the trial, 143 patients were randomized to preoperative RT then surgery (control) or preoperative pembrolizumab with RT then surgery and postoperative pembrolizumab (experimental). Remarkably, pembrolizumab improved the 2-year disease-free survival (DFS) by 15% from 52% in the control arm to 67% in the experimental arm, mainly driven by a reduction in distant disease recurrence. However, pembrolizumab increased the risk of grade 3 adverse events from 31% to 56%, and most patients did not benefit from pembrolizumab. Improvement in DFS was predominantly driven by patients with grade 3 disease, but additional biomarkers will be critical to identify the patients with stage III STS most likely to benefit from pembrolizumab.

Due to their mesenchymal origin, STS cells form complex interactions with other stromal cells in the tumor microenvironment (TME), which might explain the poor performance of PD-L1 expression and tumor mutational burden for predicting ICI response in patients with STS^13^. Several prior studies have demonstrated that the sarcoma TME has prognostic value and can potentially predict response to ICI^14,15^. For example, a previous study estimated the abundance of nine stromal cell populations by deconvoluting bulk gene expression data to define five sarcoma immune classes (SICs)^14,16^. Of these, SIC E, characterized by an inflamed TME with abundant B-cells and tertiary lymphoid structures (TLS), was associated with improved outcomes in patients with metastatic STS receiving pembrolizumab. We previously used a machine learning method called EcoTyper^17^ to identify three STS-specific cellular communities of co-occurring transcriptional cell states (sarcoma ecotypes, SEs) that were associated with prognosis and predicted response to immunotherapy across independent cohorts in metastatic STS^15^. However, it is unclear if these SICs and SEs can identify patients with localized sarcomas who could benefit from the addition of perioperative ICI to preoperative RT.

Here, we perform a comprehensive multi-omics analysis of samples from patients enrolled in the SU2C-SARC032 trial to elucidate sarcoma TME dynamics in response to RT with or without ICI and to identify predictors of benefit from pembrolizumab. We identify two distinct TME signatures associated with better outcomes after pembrolizumab and provide evidence that RT promotes intratumoral inflammation that may synergize with ICI to trigger anti-tumor immune responses in immune cold STS. If validated in future studies, our findings could help to identify patients with localized STS most likely to benefit from pembrolizumab to prevent the development of metastatic disease.

## Results

### Correlative multi-omics analysis of SU2C-SARC032 patient samples

Patients in the SU2C-SARC32 trial were enrolled between November 28, 2017 and November 14, 2023 and randomized to neoadjuvant RT (50 Gy in 25 fractions) followed by surgical resection (control) or neoadjuvant RT with pre- and postoperative pembrolizumab (experimental, **Fig. 1**). A total of 143 patients were enrolled in the trial, and 127 evaluable patients were included in the modified intention-to-treat analysis (**Supplementary Fig. S1a**). Three evaluable patients did not undergo surgery, two had disease progression before surgery, and one patient on the experimental arm discontinued treatment due to adverse events. No evaluable patients received adjuvant chemotherapy. Formalin-fixed paraffin-embedded (FFPE) pre-treatment tumor biopsies and surgical resection specimens were analyzed with bulk RNA-sequencing (RNA-seq). Overall, bulk RNA-seq data was available for 119 of 127 (94%) evaluable patients with 70/119 (59%) patients having both pre-treatment biopsy and surgical resection RNA-seq data. RNA-seq was available only for the pre-treatment time point in 41/119 (34%) and only on the surgical resection sample in 8/119 (7%). Fresh post-treatment surgical resection samples were obtained for 56/127 (44%) patients and processed through flow cytometry. Peripheral blood mononuclear cells (PBMC) samples were collected longitudinally for Cytometry by Time of Flight (CyTOF) analysis in 55 patients before starting treatment (“pre-treatment”), one week after starting treatment (“1 week on-treatment”), during the 3^rd^ week of RT (“Week 3 of RT”), after neoadjuvant treatment and before surgery (“Post-RT/Pre-Surgery”), and at 3 and 12 months after surgery. Follow up data was updated on February 24, 2025 for the correlative analyses.

**Fig. 1:**
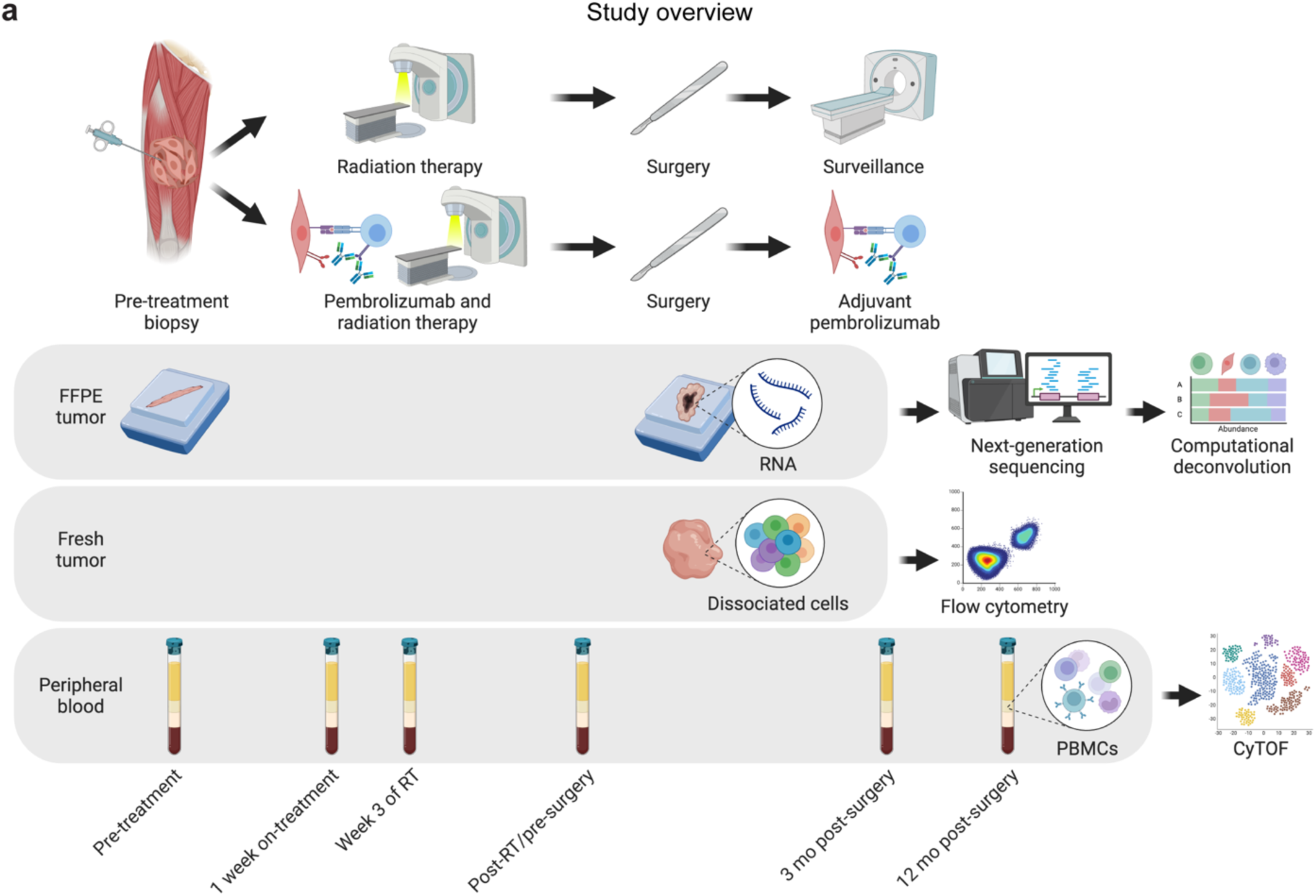
Overview of multi-omics tumor and peripheral blood analyses of patients treated in SU2C-SARC032. **a**, Patients were randomized to neoadjuvant radiotherapy followed by surgery (control) or 3 cycles of neoadjuvant pembrolizumab and radiotherapy followed by surgery and up to 14 cycles of adjuvant pembrolizumab (experimental). Bulk RNA was isolated from formalin-fixed paraffin embedded (FFPE) pre-treatment tumor biopsies and surgical resection tumor specimens prior to next-generation sequencing and digital cytometry to measure cell state abundances. Fresh surgical resection specimens after preoperative radiotherapy with or without pembrolizumab were dissociated prior to analysis with flow cytometry. Peripheral blood mononuclear cells (PBMCs) were isolated from longitudinal peripheral blood samples for analysis using cytometry by time-of-flight (CyTOF).

### Baseline sarcoma TME signatures correlate with outcomes

Based on the established association between SICs and SEs with patient outcomes in localized STS and response to ICI in patients with metastatic STS^14,15^, we investigated if these STS TME signatures were associated with outcomes in patients enrolled in SU2C-SARC032. We assigned each patient to an SIC based on bulk RNA-seq of their pre-treatment sample (**Fig. 2a**, **Supplementary Table S1**). Consistent with the prior report^14^, patients classified at baseline as SIC E, which is characterized by a rich pro-inflammatory immune infiltrate, had better DFS than patients classified as any other SIC (hazard ratio [HR]: 0.36; 95% confidence interval [CI] 0.19, 0.67; nominal *P* = 0.01, **Fig. 2b**, **Supplementary Fig. S2a**). Additionally, when considering only SIC E patients, those receiving pembrolizumab had better DFS compared to those receiving RT alone (HR: 0.37; 95% CI 0.07, 1.85; nominal *P* = 0.23, **Fig. 2c**). In contrast, among patients belonging to any other SIC than SIC E, there was little difference in DFS between those in the control versus experimental arm (HR: 0.86; 95% CI 0.47, 1.55; nominal *P* = 0.59, **Fig. 2d**).

**Fig. 2:**
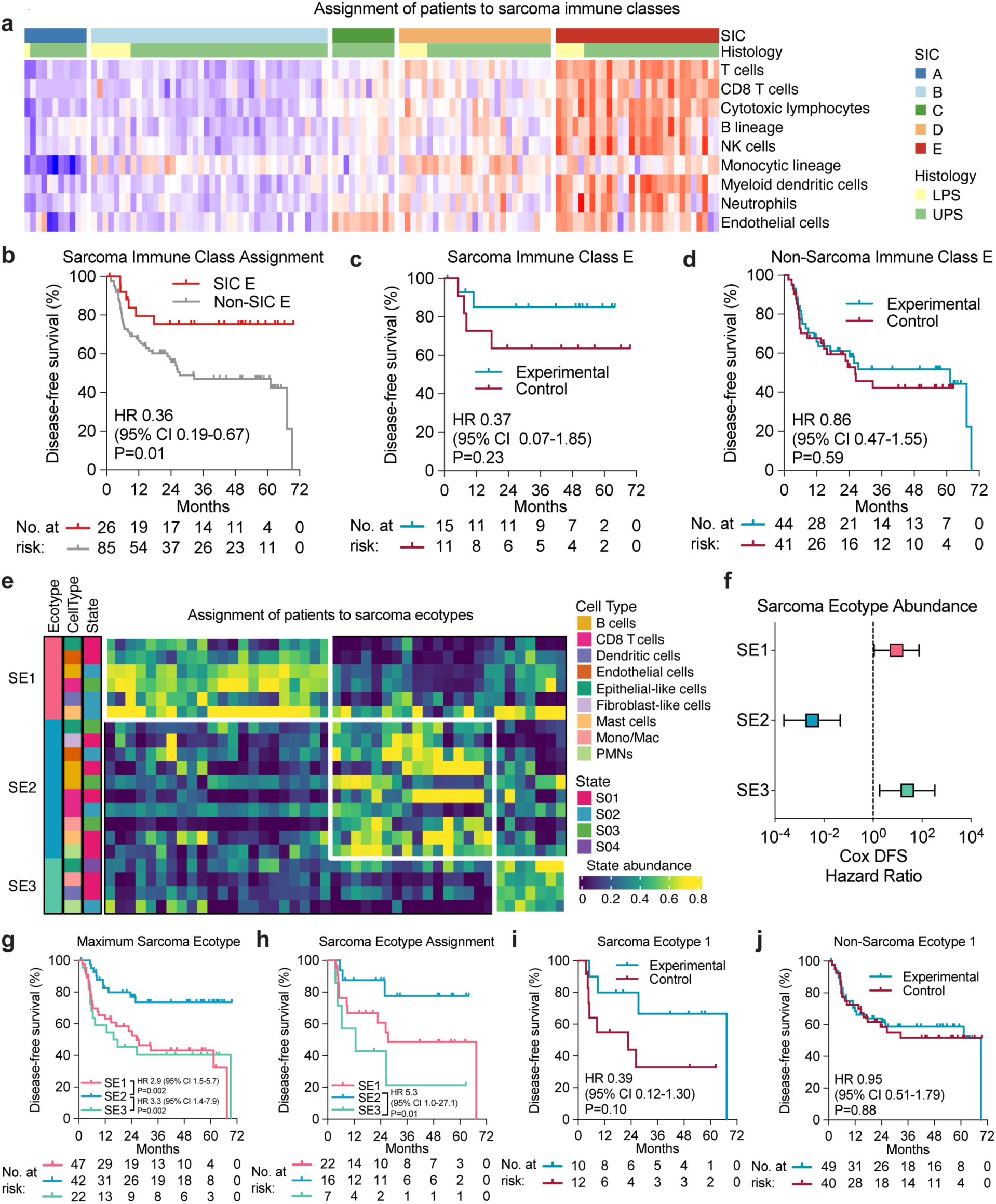
Association of pre-treatment sarcoma microenvironment signatures with outcomes. **a**, Heat map of cell type abundances in the tumor microenvironment of pre-treatment sarcomas measured using MCP-counter. Samples are grouped by sarcoma immune class (SIC) assignment. **b**-**d**, Kaplan-Meier curves showing disease-free survival (DFS) for (**b**) evaluable patients in SIC E versus all other SICs, (**c**) SIC E patients in the experimental versus control arm, and (**d**) non-SIC E patients in the experimental versus control arm. **e**, Heat map of cell state abundances in the tumor microenvironment of pre-treatment sarcomas measured using EcoTyper. Samples are grouped by sarcoma ecotype (SE) assignment. **f**, Cox proportional hazard ratios for DFS based on SE abundance. **g-j**, Kaplan-Meier curves showing DFS for evaluable patients based on (**g**) maximum SE, (**h**) SE assignment, (**i**) patients assigned to SE1 patients in the experimental versus control arm, and (**j**) patients not assigned to SE1 patients in the experimental versus control arm. P-values were calculated using two-sided log-rank tests.

Next, we applied the EcoTyper framework to measure the abundance of the three previously defined SEs based on the pre-treatment tumor RNA expression profile (**Fig. 2e**). When analyzing SE abundance as a continuous variable (**Fig. 2f**), analyzing patients based on the SE with the maximum abundance (**Fig. 2g**), or assigning patients to a dominant SE when present (**Fig. 2h**), SE2 was associated with better DFS than SE1 and SE3, consistent with our prior report^15^. This is consistent with SE2 tumors being enriched for activated pro-inflammatory immune cell states. In contrast, SE3 tumors are enriched for immunosuppressive cell states such as *SPP1*^+^ M2-like tumor-associated macrophages, and SE1 tumors have low proportions of immune cells overall. Interestingly, despite being immune cold tumors with low response rates to ICI alone in the metastatic setting^15^, patients assigned to SE1 that received pembrolizumab appeared to have better DFS than those receiving RT only (HR: 0.39; 95% CI 0.12, 1.3; nominal *P* = 0.10, **Fig. 2i**). There was minimal difference in DFS between non-SE1 patients treated with pembrolizumab and those treated with RT only (HR: 0.95; 95% CI 0.51, 1.79; nominal *P* = 0.88, **Fig. 2j**). Because patients with grade 3 tumors appeared to derive most of the benefit from pembrolizumab in SU2C-SARC032^12^, we evaluated whether there was a difference in the proportion of grade 2 and 3 tumors based on SIC or SE classification. Grade 2 tumors were more abundant in SIC A compared to SIC B patients (**Fig. S2b**, 60% versus 23%, respectively, nominal *P* = 0.049) and SE2 compared to SE3 patients (**Fig. S2c**, 56% versus 0%, respectively, nominal *P* = 0.02). However, neither SIC E nor SE1 were enriched for grade 3 versus grade 2 tumors.

Next, we asked if the SIC and SE classifications captured separate or complementary biological features of the sarcoma TME. Consistent with SE1 being immune cold and SE2 being immune hot, the median SE1 abundance was highest in SIC A patients (the most immune cold SIC) while SE2 abundance was highest in SIC E and SIC D (the most immune hot SICs, **Fig. S2d**). Similarly, the proportion of patients classified as SE1 based on maximum abundance decreased from SIC A to SIC E while the proportion of patients classified as SE2 increased (**Fig. S2e, Fig. S2f**). Of the 10 patients classified as SIC A, all had SE1 as the most abundant SE. However, patients in the other SICs had a mixture of all three SEs as the most abundant SE, demonstrating the complementarity of these TME signatures.

Next, we measured the expression of 18 genes comprising a T cell-inflamed gene expression profile (GEP), which was previously associated with response to pembrolizumab in patients across 9 different cancers^18^ (**Supplementary Table S2**). The pre-treatment T cell-inflamed GEP score was associated with longer DFS in patients that received pembrolizumab (HR: 0.62; 95% CI 0.4, 0.6; nominal *P* = 0.02, **Fig. S2g**). Interestingly, the T cell-inflamed GEP score was highest in patients belonging to SIC E (mean T cell-inflamed GEP: SIC E 4.7, SIC D 4.11, SIC C 3.3, SIC B 2.9, SIC A 1.9; nominal *P* < 0.0001, **Fig. S2h**), but the T cell-inflamed GEP score was lowest for patients classified as SE1 compared to SE2 or SE3 (mean T cell-inflamed GEP: SE1 2.5, SE2 4.7, SE3 3.9; nominal *P* < 0.0001, **Fig. S2i**). Overall, these findings confirm the association of SICs and SEs with prognosis in high-risk, localized STS and suggest that two TME signatures (immune hot SIC E and immune cold SE1) identify different subsets of patients who benefit from the addition of perioperative pembrolizumab to preoperative RT and surgery.

### Digital cytometry dissects pre-treatment STS TME cell states

To further dissect the cell states contributing to the STS TME before and after treatment, we performed digital cytometry on the bulk gene expression obtained from the FFPE tumor samples. We built a scRNA-seq atlas of UPS and LPS samples^19^, identifying different malignant and non-malignant TME cell states across 65,786 single cells from 12 patients (**Fig. 5a**). Overall, we identified 9 malignant cell states, 7 non-malignant mesenchymal cell states including cancer-associated fibroblasts (CAFs) and endothelial cells, 17 lymphoid, and 12 myeloid immune cell states (**Fig. S3a**, **Supplementary Tables S3** and **S4**). We derived a single-cell gene expression signature matrix from the atlas to deconvolute the abundance of each cell state in bulk RNA-seq samples with CIBERSORTx (**Supplementary Tables S5**, **S6**, and **S7**)^20^. To validate the accuracy of our deconvolution strategy, we correlated the abundance of major immune cell types in post-treatment tumor samples measured with CIBERSORTx versus flow cytometry and observed correlation despite the different tumor regions analyzed and technical approaches utilized (**Fig. S3b**).

Next, we asked if the abundance of the recovered CIBERSORTx cell states reflected the underlying biology of SEs and SICs. When analyzing cell states correlated with SE1 abundance or with higher abundance in patients assigned to SE1, we found enrichment of multiple sarcoma cell states (**Fig. 3b**, **Fig. 3c, Fig. S4a**), including mesenchymal-like ECM remodeling sarcoma cells with high expression of *MMP2*, *COL6A3*, and *COL1A2* and proangiogenic STS cell states with high expression of hypoxia-related genes (*CA9*, *CA12*) and proangiogenic mediators (*VEGFA1*, *ANGPTL4*). Interestingly, non-tumoral cell states with similar functionality, including matrix CAFs associated with extra-cellular matrix (ECM) remodeling^21^ and vascular CAFs implicated in angiogenesis were also enriched in SE1 tumors. Accordingly, capillary venous ECs, Tip ECs, arterial ECs, and pericytes were all elevated in SE1 tumors, suggesting that pro-angiogenic, pro-fibrotic, stromal and tumoral niches dominate their TME. Most myeloid and lymphoid immune cell states were negatively correlated with SE1 abundance, with the notable exception of CD1c^+^ dendritic cells, which are involved in antigen presentation^22^, and M2 macrophages with high expression of *APOE* and *APOC1*, which have been associated with resistance to immune checkpoint inhibition^23^. When analyzing the abundance of the CIBERSORTx cell states in tumors assigned to each SIC, we observed enrichment in pro-inflammatory immune cell states in SIC E compared to other SICs, including B cells and plasma cells, activated CD8^+^ T cells, *GZMK*^+^ CD8^+^ T memory cells, and CD56^low^ NK cells (**Fig. 3d**, **Fig. S4b**). We also observed enrichment of myeloid cell subsets, like amphiregulin-expressing dendritic cells (*AREG*^+^ DCs)^24^ and *SELENOP*^+^ *SLC40A1*^+^ macrophages, and exhausted CD8^+^ T cells in SIC E samples, possibly indicating the establishment of a chronic anti-tumor immune response leading to T cell dysfunction^25,26^.

**Fig. 3:**
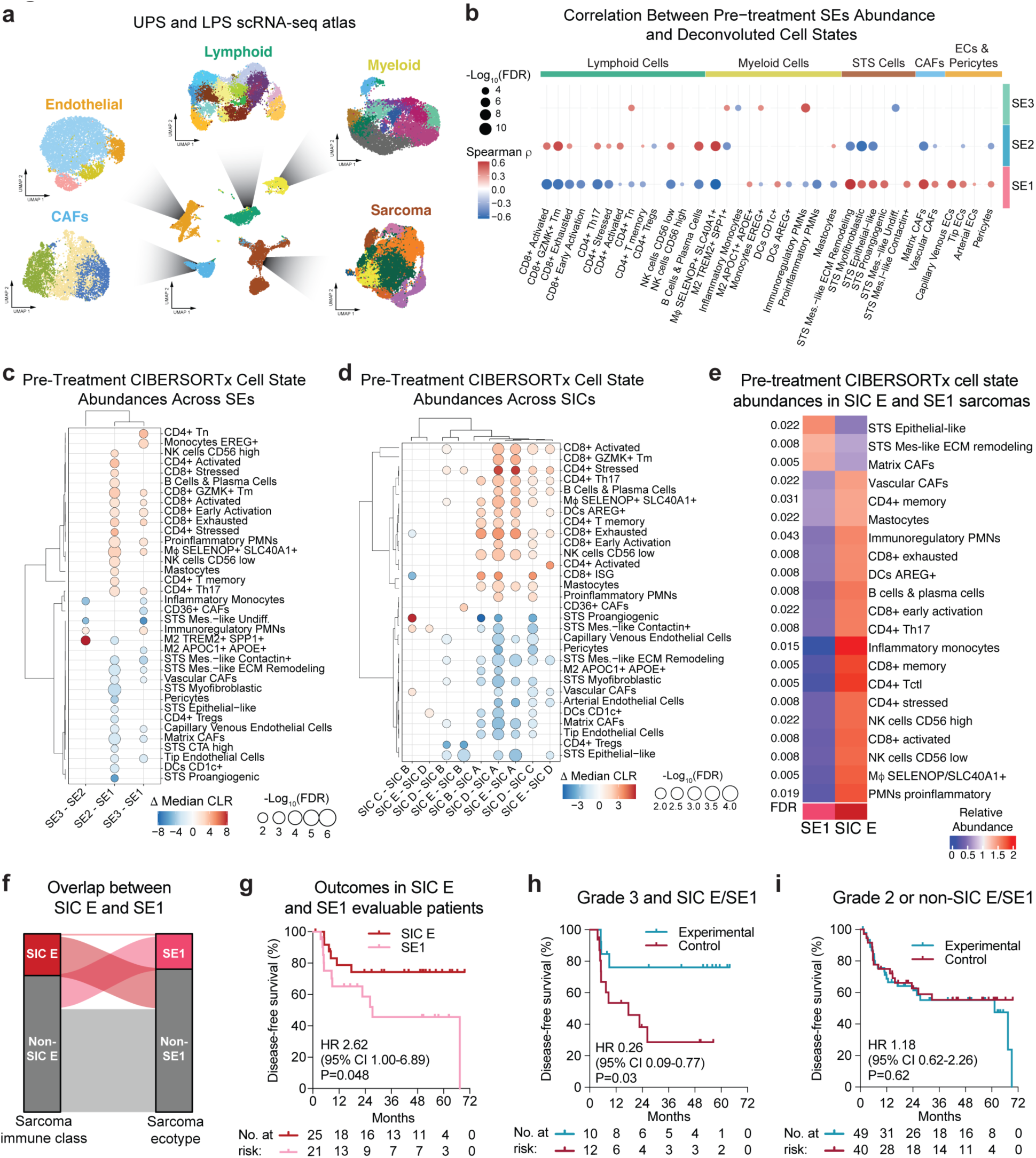
Digital cytometry of the pre-treatment sarcoma microenvironment and an integrated predictor of benefit from pembrolizumab. **a**, UMAPs showing major cell type clusters and cell state subclusters defined from single cell RNA-sequencing (scRNA-seq) of undifferentiated pleomorphic sarcomas (UPS) and liposarcomas (LPS). This scRNA-seq atlas was used to create a signature matrix for digital cytometry of bulk RNA-seq data with CIBERSORTx. **b**, Dot plot showing spearman correlation between CIBERSORTx cell state abundance and sarcoma ecotype (SE) abundances in the pre-treatment samples. The dots are colored using the spearman correlation coefficient ρ, while the dot size represents the - Log_10_(FDR) obtained using the Benjamini-Hochberg method for multiple hypothesis testing correction. Correlations with an FDR < 0.05 are shown. **c**, Dot plot showing difference in median centered log ratio (CLR) abundance of pre-treatment CIBERSORTx cell states based on pre-treatment SE assignment. Only cell states with FDR < 0.05 on Kruskal-Wallis test and pairwise SIC comparisons with a Dunn’s test FDR < 0.05 are shown. **d**, Dot plot showing difference in median centered log ratio (CLR) abundance of pre-treatment CIBERSORTx cell states based on sarcoma immune class (SIC) assignment. Only cell states with FDR < 0.05 on Kruskal-Wallis test and pairwise SIC comparisons with a Dunn’s test FDR < 0.05 are shown. **e**, Heat map showing CIBERSORTx cell states with different pre-treatment abundances between SIC E and SE1 sarcomas. P-values were calculated using two-sided Wilcoxon rank-sum tests. FDRs were obtained by correcting P-values for multiple hypothesis testing using the Benjamini-Hochberg method. **f**, River plot showing overlap between sarcoma immune class E (SIC E) and sarcoma ecotype 1 (SE1). The thickness of the bars represents the proportion of patients in each group. **g**, Kaplan-Meier curves showing the disease-free survival for evaluable patients assigned to SIC E versus SE1 based on pre-treatment RNA-sequencing. P-value was calculated using a two-sided log-rank test. **h** and **i**, Kaplan-Meier curves showing disease-free survival by treatment arm for (**h**) patients with grade 3 tumors assigned to SIC E or SE1 and (**i**) patients with grade 2 tumors or tumors not assigned to SIC E or SE1. P-values were calculated using a two-sided log-rank tests. M<Ι: macrophages.

When directly comparing the abundance of the deconvoluted cell states between SIC E and SE1, we found that SIC E tumors were enriched for both activated and stressed/exhausted CD4^+^ and CD8^+^ T cell states, B cells and plasma cells, and CD56^high^ and CD56^low^ NK cells (**Fig. 3e**). In contrast, SE1 sarcomas were enriched for both epithelial-like and mesenchymal-like ECM remodeling sarcoma cell states and matrix CAFs. Further demonstrating the difference between these two TMEs, only one patient was classified as both SE1 and SIC E at baseline (**Fig. 3f**), and SE1 patients had worse DFS compared to SIC E patients when considering all evaluable patients across both treatment arms (HR: 2.62; 95% CI 1.0, 6.9; nominal *P* = 0.048, **Fig. 3g**).

Given the observed benefit of pembrolizumab in SU2C-SARC032 for patients with grade 3 disease^12^, we asked if combining tumor grade with baseline SIC and SE classification could better identify patients that would benefit the most from pembrolizumab. Interestingly, we found that patients classified as either SIC E or SE1 that also had grade 3 tumors showed a DFS benefit from the addition of pembrolizumab (**Fig. 3h**, HR: 0.26; 95% CI 0.09, 0.8; nominal *P* = 0.03), while those with grade 2 disease or classified as non-SIC E and non-SE1 did not appear to benefit from immunotherapy (**Fig. 3i**, HR: 1.18; 95% CI 0.6, 2.3; nominal *P* = 0.62, interaction *P* = 0.03). Overall, these results highlight the differences between SE1 and SIC E tumors and suggest that integrating these complementary TME signatures with tumor grade can help to identify most patients who benefit from pembrolizumab in SU2C-SARC032.

### Neoadjuvant RT plus ICI reshapes the sarcoma TME

To define the effects of neoadjuvant RT with or without ICI on the STS TME, we performed differential gene expression and gene set enrichment analysis on the pre-treatment and post-treatment samples from both arms. When accounting for the effect of pembrolizumab, post-RT samples showed enrichment for gene sets associated with an inflammatory response, including IFN-γ, IL-2, and IL-6 signaling, as well as gene sets associated with anti-tumor responses like allograft rejection and apoptosis (**Fig. 4a**). In patients treated with pembrolizumab, we observed a down-regulation of genes associated with epithelial to mesenchymal transition and MYC targets and an enrichment for genes associated with normal myogenesis, which might reflect reduced proliferation and decreased abundance of cancer cells (**Fig. 4b**). Next, we measured the pre- and post-treatment T cell-inflamed GEP score in patients in the control and experimental arms (**Fig. S5a**, **S5b**) and found a post-treatment increase in the T cell-inflamed GEP score only in baseline SE1 patients receiving pembrolizumab plus RT (**Fig. S5c**). Similar results were observed when patients were grouped by SIC, where there was a more pronounced increase in the T cell-inflamed GEP score post-treatment for SIC A or SIC B tumors and mainly among patients receiving pembrolizumab (**Fig. S5d**). To test if these differences in immune activation profiles also corresponded to differences in the T cell repertoire, we reconstructed the T cell receptor (TCR) sequences from pre- and post-treatment bulk RNA-seq samples using TRUST4^27^ (**Fig. 4c**). Only in patients receiving pembrolizumab and RT was there a post-treatment increase in TCR repertoire diversity (Shannon entropy, q = 0.0004; Inverse Simpson index, q = 0. 0004) and clonotype richness (q = 0. 0004). These results suggest increased recruitment and expansion of multiple T cell clones in response to the combination of RT and pembrolizumab as opposed to RT alone.

**Fig. 4:**
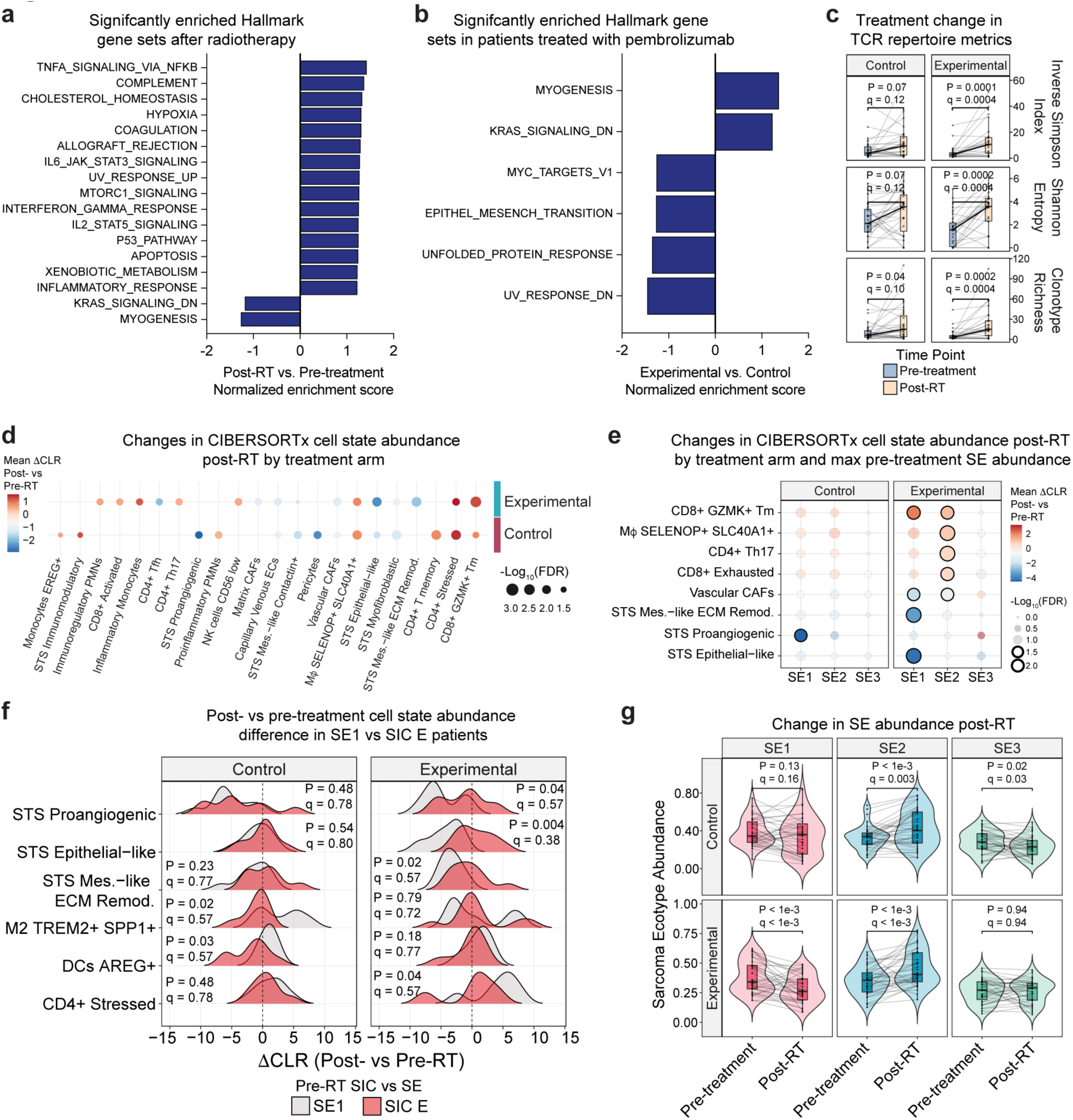
Effect of radiotherapy with or without pembrolizumab on the sarcoma microenvironment. **a**-**b**, Bar plots showing gene set enrichment analysis (GSEA) results comparing (**a**) post-RT versus pre-treatment samples accounting for the effect of pembrolizumab and (**b**) experimental versus control samples measured as the interaction of pembrolizumab with radiotherapy. **c**, Box plots showing the post-RT change in T-cell receptor repertoire metrics recovered with TRUST4 in the control and experimental arms. P-values were calculated using two-sided Wilcoxon signed-rank tests and adjusted for multiple hypothesis testing using the Benjamini-Hochberg method. **d**, Dot plot showing difference in mean centered log ratio (CLR) abundance of CIBERSORTx cell states between pre- and post-treatment samples in the control and experimental arms. P-values were calculated using a Wilcoxon signed-rank test and adjusted for multiple hypothesis testing using the Benjamini-Hochberg method. Only cell states with FDR < 0.05 are shown. **e**, Dot plot showing difference in mean CLR abundance of CIBERSORTx cell states between pre- and post-treatment samples in the control and experimental arm. Patients are group based maximum pre-treatment SE abundance. P-values were calculated using a Wilcoxon signed-rank test and adjusted for multiple hypothesis testing using the Benjamini-Hochberg method. **f**, Ridge plot showing distribution of the post- vs pre-RT difference in CLR abundance of CIBERSORTx cell states between patients classified as SE1 and those classified as SIC E at baseline. P-values were obtained using a two-sided Wilcoxon rank-sum test and adjusted for multiple hypothesis testing using the Benjamini-Hochberg method. **g**, Box-violin plots showing the abundance of sarcoma ecotypes before and after treatment in patients in the experimental and control arms. P-values were computed using Wilcoxon signed-rank tests and corrected for multiple comparisons using the Benjamini-Hochberg method. M<Ι: macrophages.

Next, we queried if the addition of pembrolizumab to RT changed the overall composition of the TME by measuring the treatment-related change in abundance of the cell states recovered through CIBERSORTx (**Fig. 4d**). Interestingly, there was a decrease in matrix CAFs and ECM remodeling mesenchymal-like sarcoma cells only in patients that received pembrolizumab and a greater decrease in epithelial-like sarcoma cells in the experimental versus the control arm. Also, CD8^+^ activated T-cells and the highly cytotoxic CD56^low^ NK cell state^28^ were both increased post-RT in the experimental arm but not in the control arm. At the same time, we observed an increase in both immunoregulatory PMNs and proinflammatory monocytes in patients receiving pembrolizumab, which might indicate the establishment of compensatory immunoregulatory mechanisms in response to ICI. In contrast, in patients that received RT only, we saw a reduction in pericytes and proangiogenic STS states, possibly a result of RT-induced tumor vascular remodeling^29^. Also, in patients receiving RT-only there was an increased abundance of immune suppressive EREG^+^ monocytes and immunomodulatory STS states, likely reflecting a compensatory anti-inflammatory response to RT with increased recruitment of myeloid-derived suppressor cells (MDSCs)^30^.

To determine if RT with or without ICI led to different changes in TME composition based on baseline SE, we measured the change in CIBERSORTx cell state abundance post-RT within the control and experimental arm based on the most abundant SE pre-treatment (**Fig. 4e**). Patients with baseline SE1 tumors receiving pembrolizumab had a higher post-treatment increase in *GZMK*^+^ CD8^+^ T memory cells and a more pronounced reduction in both vascular CAFs and epithelial-like and mesenchymal-like STS cell states than baseline SE1 tumors treated with RT alone. Given the observed benefit from the addition of ICI to RT in both patients with baseline SE1 and SIC E tumors, we asked if there was a divergent pattern in the post-treatment changes in CIBERSORTx cell states between patients classified as SIC E and SE1 at baseline (**Fig. 4f**). We observed a trend towards a more pronounced post-treatment decrease in both mesenchymal-like, epithelial-like, and proangiogenic STS cell states and an increase in stressed CD4 T cells in SE1 patients compared to SIC E patients in the experimental arm. Lastly, we observed a marked decrease in the immune cold SE1 abundance in patients that received pembrolizumab but not in those treated with RT alone and an increase in the immune hot SE2 that was more pronounced in the experimental compared to the control arm (**Fig. 4g**, **Supplementary Table S8**). Taken together, these results support a model where the addition of pembrolizumab to neoadjuvant RT induces immune activation in immune cold SE1 sarcoma while also reigniting an established yet dysfunctional pre-existing anti-tumor response in immune hot SIC E sarcomas.

### Longitudinal systemic immune monitoring through CyTOF

Peripheral blood immune cell profiling has been shown to correlate with response to immunotherapy across different cancers in the metastatic setting^31^. Based on this, we asked if neoadjuvant RT with or without pembrolizumab could result in systemic immune changes detectable in the peripheral blood of STS patients using CyTOF analysis of PBMCs (**Supplementary Tables S9**, **S10**, and **S11**). Unsupervised clustering revealed 9 major PBMC clusters including B cells, monocytes, CD4^+^ T cells, naïve CD8^+^ T cells, effector CD8^+^ T cells, activated T cells, memory T cells, NK cells, and CD56^+^ NK cells, and further sub-clustering of peripheral blood T cells revealed 11 sub-populations (**Fig. 5a**, **5b**, **5c**, and **5d**).

**Fig. 5:**
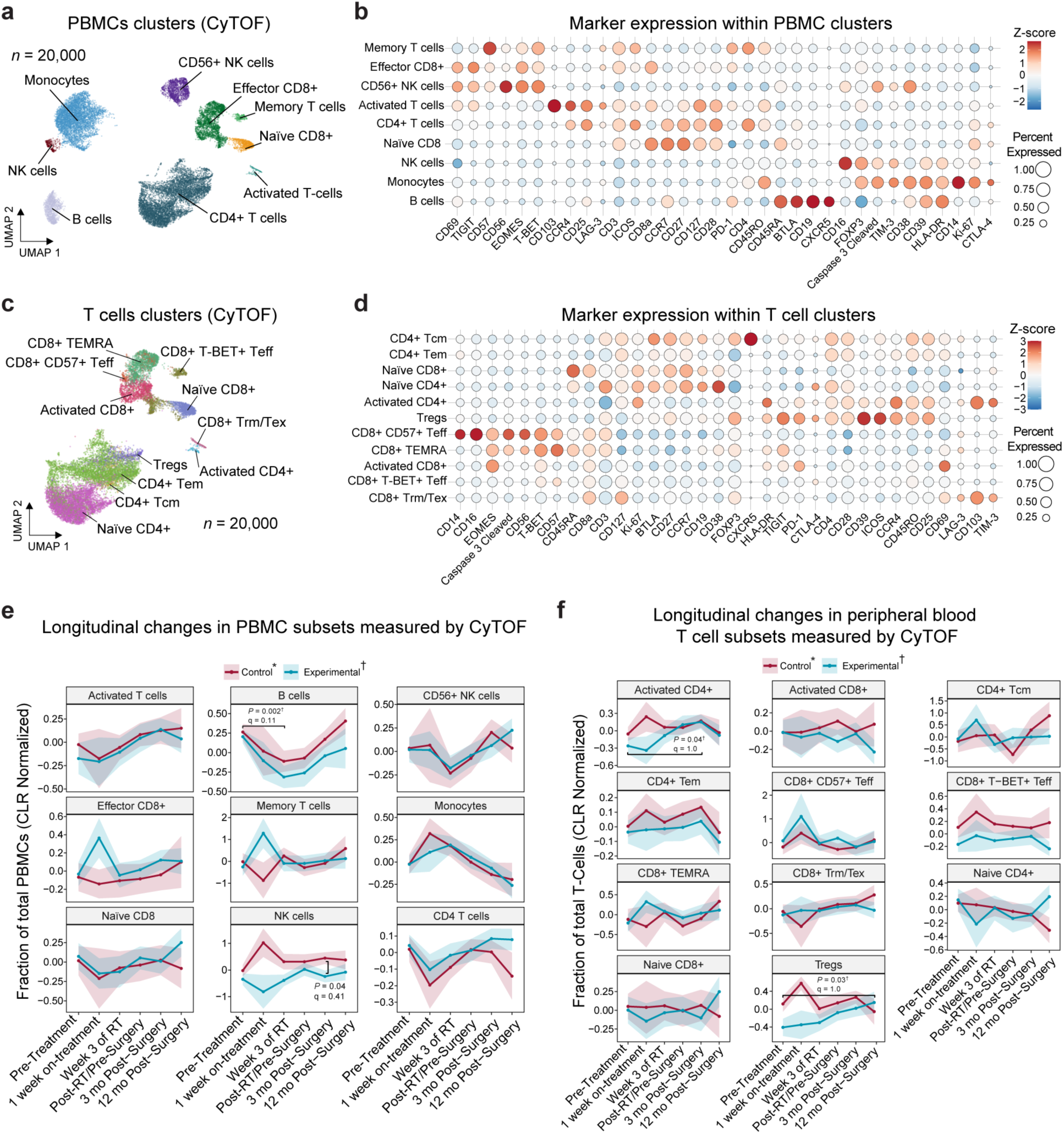
Longitudinal cytometry by time-of-flight (CyTOF) analysis of peripheral blood mononuclear cells (PMBCs). **a**, Unsupervised clustering of PBMCs based on surface marker expression measured by CyTOF. 20,000 cells were randomly selected for plotting. **b**, Dot plot showing expression of surface markers in PBMCs clusters. **c**, Unsupervised clustering of T cell subsets based on surface marker expression measured by CyTOF. 20,000 cells were randomly selected for plotting. **d**, Dot plot showing expression of surface markers in T cell clusters. **e**-**f**, Longitudinal changes in (**e**) PMBC subsets and (**f**) peripheral blood T cell subsets over time in the experimental and control arms. Plotted values represent centered log-ratio (CLR) normalized fractions of total PBMCs or T cells. P-values were calculated using two-sided Wilcoxon rank-sum tests. Multiple hypothesis testing correction was applied using the Benjamini-Hochberg method (q-value).

Considering the PBMC clusters (**Fig. 5e**), there was a reduction in the abundance of B cells between the pre-treatment time point and the 3^rd^ week of RT in both the control arm (mean centered log-ratio [CLR] 0.26 vs −0.11, nominal *P* = 0.002, q = 0.11) and the experimental arm (mean CLR 0.20 vs −0.31, nominal *P* = 0.03, q = 0.78). In contrast, the abundance of memory T cells tended to increase at 1 week on-treatment in patients in the RT plus pembrolizumab arm (mean CLR −0.25 vs 1.28, nominal *P* = 0.06, q = 0.91) as opposed to patients that only received RT (mean CLR −0.01 vs −0.87, nominal *P* = 0.29, q = 0.81). Lastly, patients that received RT only compared to those that also received pembrolizumab had higher levels of circulating NK cells at 3 months post-surgery (mean CLR −0.03 vs −0.09, nominal *P* = 0.04, q = 0.41). Considering peripheral blood T cell subsets (**Fig. 5f**), activated CD4+ T cells increased at 3 months post-surgery for patients that received pembrolizumab (mean CLR −0.26 vs 0.15, nominal *P* = 0.02, q = 1.0), while circulating T-regs tended to increase at one year post-surgery compared to before starting treatment in patients in the experimental arm (mean CLR 0.14 vs −0.40, nominal *P* = 0.03, q = 1.0). However, most PBMC subsets did not substantially change over the course of treatment. Overall, these results indicate modest post-treatment changes in peripheral blood immune cell subsets in patients with STS receiving immunotherapy and RT.

### Flow cytometry reveals post-treatment TME architecture

For patients with viable fresh tumor specimens collected after neoadjuvant therapy at the time of surgical resection, we measured treatment effect on tumor immune cell infiltration using flow cytometry (**Supplementary Tables S12** and **S13**). Among CD4^+^ T cells, we observed a lower proportion of CD45RA^+^ CCR7^+^ naïve cells (mean ΔCLR: −0.51, q = 0.03) and a higher proportion of CD45RA^−^ CCR7^−^ effector memory (Tem) cells (mean ΔCLR: 0.50, q = 0.03) in patients that received neoadjuvant pembrolizumab and RT compared to those that received RT only (**Fig. 6a**, **6b**, and **6c**), consistent with a switch towards an effector T cell phenotype. Similar changes were observed within the CD8^+^ T cell population, though these were less marked. Contrasting patients classified as SE1 versus SIC E at baseline, central memory T cells decreased in SE1 patients but increased in SIC E patients treated with pembrolizumab. In addition, we observed a more pronounced increase in effector memory T cells and a more pronounced decrease in naïve T cells for SE1 patients treated on the experimental arm.

**Fig. 6:**
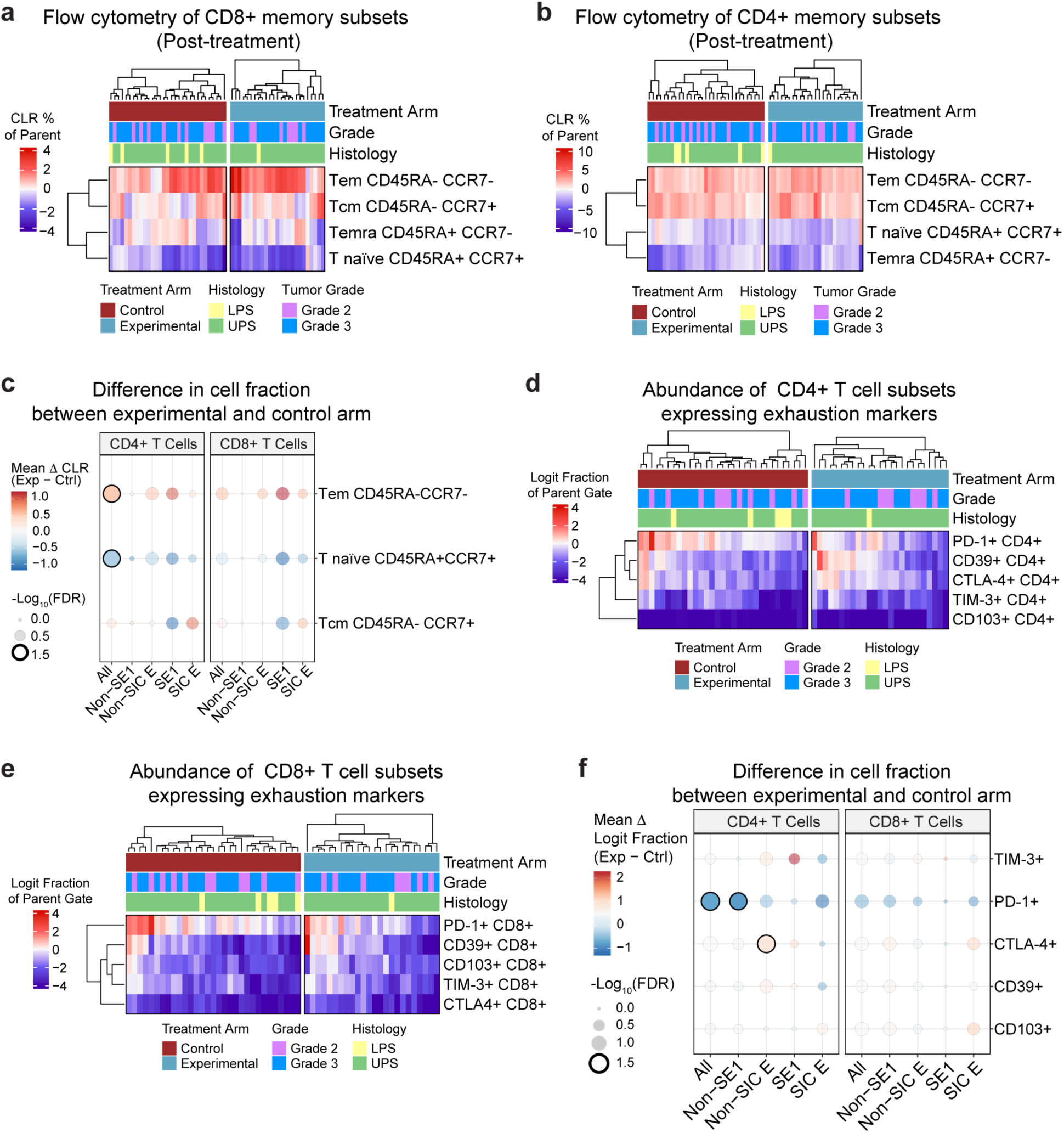
Flow cytometry analysis of post-treatment samples. **a** and **b**, Heat maps showing the abundance of (**a**) CD8+ T cell, and (**b**) CD4+ T cell memory subsets in resected sarcomas after radiotherapy with or without pembrolizumab measured by flow cytometry. Patient samples are grouped according to study arm. Cell type abundances are reported as CLR normalized fractions of the total number of cells in the parent gate. **c**, Dot plot showing mean difference in CD4+ and CD8+ memory subsets CLR abundance between patients in the experimental and control arm. Samples are grouped based on baseline SE1 and SIC E classification. P-values were calculated using two-sided Wilcoxon rank-sum tests. Multiple hypothesis testing correction was applied using the Benjamini-Hochberg method. **d** and **e**, Heat maps showing the abundance of (**d**) CD4+ T cell, and (**e**) CD8+ T cell subsets expressing different exhaustion markers in resected sarcomas after radiotherapy with or without pembrolizumab measured by flow cytometry. Patient samples are grouped according to study arm. Cell type abundances are reported as logit normalized fractions of the total number of cells in the parent gate. **f**, Dot plot showing mean difference in abundance of CD4+ and CD8+ T cell subsets expressing different exhaustion markers, measured as logit fraction of parent gate, between patients in the experimental and control arm. Samples are grouped based on baseline SE1 and SIC E classification. P-values were calculated using two-sided Wilcoxon rank-sum tests. Multiple hypothesis testing correction was applied using the Benjamini-Hochberg method.

When analyzing activation and exhaustion markers in CD4^+^, CD8^+^, and regulatory T cells, we observed a lower proportion of PD-1^+^ cells in patients in the experimental arm versus the control arm (**Fig. 6d**, **6e**, **6f, S7a,** and **S7b**), which might be explained by antigen masking by pembrolizumab. There was a higher proportion of TIM-3^+^ CD4^+^ T-cells (mean Δ logit fraction: 2.2, q = 0.27) in SE1 patients receiving pembrolizumab, while the opposite trend was noted in SIC E patients. Among myeloid cells, pembrolizumab increased the proportion of CD68^+^ CD163^+^ M2-like macrophages (mean Δ logit fraction: 0.89, q = 0.18, **Fig. S7c**, **S7d**) and CD14^+^CD16^-^ classical monocytes (mean Δ logit fraction: 0.75, q = 0.07, **Fig. S7e**, **S7f**) in SE1 patients with little change in SIC E patients. Overall, these results indicate that adding pembrolizumab to neoadjuvant RT results in an overall shift of tumor infiltrating lymphocytes towards an effector phenotype that was most pronounced in baseline immune cold SE1 tumors.

## Discussion

In this study, we performed a comprehensive multi-omics assessment of patients with stage III LPS or UPS of the extremity receiving neoadjuvant RT with or without perioperative pembrolizumab as part of SU2C-SARC032^12^. Our analyses revealed two distinct baseline STS TME states associated with benefit from the addition of pembrolizumab to neoadjuvant RT. We found that both an immune desert state, corresponding to SE1, and an immune hot TME, represented by SIC E, correlated with clinical benefit from the addition of pembrolizumab to RT. This observation indicates complementary mechanisms of benefit from ICI and RT^32^.

Similar to other solid tumors that respond to ICI^33^, the presence of an established anti-tumor immune response in SIC E STSs was associated with benefit from the addition of immunotherapy to RT. SIC E tumors are characterized by high expression of gene signatures related to CD8^+^ and CD4^+^ T cells, NK cells, and B cells and are enriched for TLS on IHC, indicating a robust chronic anti-tumor response^14^. Our deconvolution strategy provides a high-resolution view of the cellular states that contribute to defining SIC E and is consistent with prior observations that SIC E STSs might be primed for benefit from anti-PD-1/PD-L1 agents^14,15^. Flow cytometry analyses on post-treatment samples in this study revealed that, in SIC E tumors, the addition of ICI to RT mainly resulted in depletion of likely exhausted PD-1^+^ CD8^+^ and PD-1^+^ CD4^+^ T cells, which could lead to a reinvigorated T cell effector function^34,35^. However, it must be considered that the observed depletion of PD-1^+^ T-cell subsets in patients that received pembrolizumab might be partially driven by binding of pembrolizumab itself to PD-1. Additionally, while the association between an SIC E TME state and benefit from ICI was previously demonstrated in the metastatic setting^14^, our results also indicate a clinical benefit from immunotherapy in localized STSs with an immune hot TME. This is also consistent with data from a recent phase 2 randomized non-comparative trial (NCT03307616), which studied neoadjuvant nivolumab or ipilimumab/nivolumab in patients with resectable retroperitoneal dedifferentiated LPS (DDLPS) and neoadjuvant nivolumab combined with RT for patients with UPS of the extremity^36^. This study showed that patients with DDLPS that received neoadjuvant ICI and showed presence of TLS at surgery had improved overall survival and that baseline immune cell infiltration, rather than PD-L1 expression, correlated with outcomes in both cohorts, which is consistent with our observations.

Our study also suggested a benefit from the addition of immunotherapy for patients with baseline SE1 tumors, which typically have very little immune infiltration, likely signaling lack of a pre-existing immune response^15^. Indeed, digital cytometry revealed a low baseline abundance of activated CD8^+^ T cells in SE1 tumors with a higher proportion of STS cell states, which is consistent with the lack of anti-tumor immunity. After pembrolizumab and RT, SE1 tumors showed an increase in CD4^+^ and CD8^+^ Tem cells by flow cytometry, as well as in increase in CD8^+^ GZMK^+^ memory T cells by digital cytometry indicating the induction of a *de novo* immune response. Accordingly, we observed a post-treatment increase in the T cell-inflamed GEP score only in baseline SE1 patients treated with the RT/ICI combination. This was accompanied by an overall increase in TCR repertoire diversity and an increase in early activated CD8^+^ T cells and CD56^low^ NK cells only in the experimental arm. Notably, SE3, which was associated with response to ICI in patients with metastatic sarcomas^15^, was not associated with benefit from pembrolizumab in SU2C-SARC032. This suggests a different mechanism of response when ICI is combined with radiotherapy or in patients with gross versus micrometastatic disease. In contrast with prior studies in metastatic cancer^43,44^, our longitudinal CyTOF analysis observed only modest changes in PBMC and T cell subsets during and following treatment, suggesting that analysis of the primary tumor may be necessary to measure the effects of radiotherapy with or without pembrolizumab in patients with localized STS. Overall, our findings support a model of synergism between RT and ICI, where RT potentially induces immunogenic cell death with release of damage associated molecular patterns and cytokines, TME inflammation, and improved antigen presentation, leading to accumulation of T cells that are activated by ICI^37–39^.

The clinical benefit of pembrolizumab in SU2C-SARC032 appeared to be driven by patients with grade 3 tumors who have previously been shown to have a higher risk of developing metastatic disease^40–42^. Our findings suggest that transcriptional biomarkers can be integrated with tumor grade to more precisely identify patients benefiting from the addition of pembrolizumab to preoperative RT in localized STS. Although this observation needs prospective validation prior to incorporation into routine clinical management, an integrated biomarker could help to balance the benefit-risk profile of ICI in the neoadjuvant setting due to the increased incidence of grade 3 adverse events observed in the experimental arm of SU2C-SARC032. Additionally, while this study only included LPS and UPS patients, our findings demonstrate benefit from RT plus ICI in immune cold SE1 tumors, which have not been shown to benefit from immunotherapy alone in the metastatic setting^15^. This suggests that it might be worthwhile to explore the synergism between RT and ICI in future studies in other high grade STSs, which are generally considered to be refractory to immunotherapy in the setting of ICI alone, such as leiomyosarcomas and synovial sarcomas^9^. Lastly, we acknowledge the limitations of our study and correlative analyses, which are retrospective and exploratory in nature. Our observations aim to be hypothesis-generating and will require validation in independent cohorts and larger prospective clinical studies. A phase 3 randomized trial (HARMONY) to validate the benefit of adding anti-PD-1 ICI to preoperative radiotherapy in grade 3 UPS/LPS has been approved by the Canadian Clinical Trials Group (CCTG) and has received funding from the Canadian Institute of Health Research (CIHR). This multi-institutional clinical trial will provide an opportunity to validate the biomarkers identified in SU2C-SARC032.

In summary, we show that the sarcoma TME carries prognostic and predictive value in patients with localized disease treated with preoperative RT and PD-1 blockade. If validated in independent cohorts and prospective trials, sarcoma TME signatures could provide a biologically relevant and sarcoma-specific biomarker to identify patients most likely to benefit from the addition of ICI to neoadjuvant RT.

## Materials and Methods

### Clinical trial design

SU2C-SARC032 was an open-label, randomized, superiority phase II clinical trial. Patients were enrolled at 20 academic institutions in Australia, Canada, Italy, and the United States of America. The full original and final amended trial protocol and statistical analysis plan with summaries of each amendment are available online in the appendix of the initial publication of the trial results^12^. The trial was conducted by Sarcoma Alliance for Research Through Collaboration (SARC) and funded by Stand Up to Cancer and Merck Sharp & Dohme. Pembrolizumab was provided by Merck Sharp & Dohme. The study design was conceived by the co-investigators and approved by Merck Sharp & Dohme. There was no patient or public involvement in the design, conduct, or reporting of the trial. The funders of the study had no role in data collection, data analysis, data interpretation, or writing of this report. SU2C-SARC032 was registered with ClinicalTrials.gov under NCT03092323 on March 27, 2017 (https://www.clinicaltrials.gov/study/NCT03092323).

Patients were eligible for enrollment if they were ≥12 years old with a resectable, non-metastatic, grade 2 or 3 UPS (including myxofibrosarcoma) or dedifferentiated/pleomorphic LPS of the extremity and an Eastern Cooperative Oncology Group performance status score of 0 or 1. Key exclusion criteria were prior therapy with an anti-PD-1, anti-PD-L1, or anti-PD-L2 agent, prior RT to the site of the sarcoma, concurrent active malignancy, active autoimmune disease or infection, diagnosis of immunodeficiency or systemic immunosuppressive therapy, and planned receipt of neoadjuvant or adjuvant chemotherapy. Full inclusion and exclusion criteria are provided in the clinical trial protocol.

Patients were randomly assigned (1:1, stratified by tumor grade) to receive either neoadjuvant RT (50 Gy in 25 fractions) followed by surgery (control group) or neoadjuvant intravenous pembrolizumab (200 mg) for three doses before, during, and after neoadjuvant RT followed by surgery and up to 14 doses of adjuvant pembrolizumab. The first dose of pembrolizumab was within 7 days of enrollment, and RT started within 14 days of enrollment for the control group or 1-14 days after the first pembrolizumab infusion for the experimental group. Randomization was coordinated by the central clinical trial office at SARC using permuted block randomization with random block sizes. The enrolling sites did not have access to the random allocation sequence. Patients, physicians, and data analysts were not blinded to group allocation.

### Patient sample collection

Written informed consent was obtained for all patient data and samples obtained and analyzed in this study. All human research in this study was conducted in accordance with the Declaration of Helsinki. The SU2C-SARC032 study protocol was reviewed and approved by SARC, Merck Sharp & Dohme, and the institutional review board at each institution participating in the trial. The correlative analyses were approved by the institutional review boards at Stanford University and Duke University.

### Bulk RNA sequencing

RNA was extracted from FFPE pre-treatment core needle biopsies and surgically resected tumors using RNAstorm FFPE RNA Extraction Kits (Cell Data Sciences). Tissue from pre-treatment core needle biopsies was provided as pre-cut 5 μm slides from the treating site, and RNA was extracted from 2-3 slides per patient. Surgically resected tumors were provided as tissue blocks, and RNA was extracted from 2-3 one mm cores obtained from the region with the highest sarcoma content based on pathologist review of hematoxylin and eosin-stained slides from the block. Libraries were prepared for sequencing using SMARTer Stranded Total RNA-Seq Kits v2-Pico Input Mammalian (Takara Bio) and sequenced with 150-bp paired-end reads on a NovaSeq 6000 (Illumina). FASTQ files were quasi-aligned using Salmon^45^.

### Definition of sarcoma ecotypes and sarcoma immune classes

SIC centroids were defined based on UPS and LPS samples from TCGA using the SIC labels provided by the authors^14^. Pre-processed bulk RNA-seq profiles from TCGA^46^ were downloaded and scaled to transcripts per million (TPM). TPM counts were log2-tranformed, then analyzed using MCP-Counter^16^ to measure the abundance of 10 tumor microenvironment cell types. MCP-Counter scores were Z-score transformed across all samples for each cell type, excluding fibroblasts. Samples were grouped by their assigned SIC, and the mean Z-score abundance for each cell population was computed to create centroids for each SIC. Pre-treatment bulk RNA-seq samples from SU2C-SARC032 were processed in the same manner and assigned to the closest centroid based on Euclidean distance. Sarcoma ecotypes were recovered and quantified using the EcoTyper reference-guided annotation framework^17^ by applying the previously learnt non-negative matrix factorization model^15^ with TPM normalized expression values as input.

### Flow cytometry

Freshly isolated post-treatment tumors were disaggregated using the Human Tumor Dissociation Kit Protocol (Miltenyi Biotec). Tumor cell suspensions were immediately viably cryopreserved and stored in vapor phase liquid nitrogen until thawing prior to flow cytometry analysis. All flow cytometry assays were performed in the Duke Immune Profiling Core (DIPC) according to established Standard Operating Procedures (SOP) and in a strictly ‘blinded’ manner. Viably cryopreserved tumor cell suspensions were thawed, washed, and enumerated using a Muse Cell Analyzer. High parameter immune profiling was then performed using a 25-channel custom flow cytometry panel to profile components of the tumor microenvironment. Cells were stained with a vital dye (Zombie Yellow Fixable Viability Kit, BioLegend) along with Fc block (BD Biosciences) for 15 minutes at room temperature in the dark. Cells were washed with FACSWash Buffer (D-PBS, ThermoFisher) containing 0.5% FBS. Cells were then stained using a Cytokine/Chemokine Reagent (CCR) staining cocktail containing saturating amounts of antibodies, washed, and subsequently stained with a surface staining cocktail containing saturating amounts of antibody. Following surface staining, cells were washed, permeabilized, and incubated with Fc block (BD Biosciences). After permeabilization, cells were washed with eBioscience 1X Permeabilization Solution (ThermoFisher), incubated with Fc block (BD Biosciences), washed, and stained with an intracellular staining cocktail. Following intracellular staining, cells were washed, fixed with D-PBS containing 1% formalin, and acquired using a calibrated and optimized BD FACSymphony A5 Cell Analyzer (BD Biosciences) within 6 hours of fixation. Compensation Beads (BD Biosciences) were stained for each monoclonal antibody and used to correct for spectral overlap prior to gating analysis. FlowJo version 10.10.0 (BD Biosciences) was used to analyze the data using a gating template. An internal PBMC batch control sample was included with each batch to detect potential batch-to-batch variation. Gating was performed according to best practices with all gates set based on marker expression of the batch control. Analysis gates were drawn for each panel as follows: 1) Time gate was used to exclude air bubbles and clogs that might have occurred during acquisition for all files, 2) Zombie Yellow Fixable Viability dye was used to exclude non-viable cells, and 3) Singlet gating of both Forward and Side Scatter was used to exclude aggregates. Given the compositional nature of cell fractions obtained through flow cytometry, where for each patient at each time point the fraction of different cell subsets sum up to 1 within a given parent gate, centered log-ratio transformation was applied on the raw fractions before downstream analysis using the “compositions“ R package^47^. For overlapping non-mutually exclusive subsets within a population, to avoid infinite values at either 0% or 100% positivity, cell fractions were logit transformed as follows:

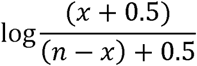

Where *x* is the number of marker positive events, and *n* is the number of events in the parent population.

### CyTOF

SU2C-SARC032 patient PBMCs collected prior to treatment, one week after starting treatment, during the 3^rd^ week of RT, after neoadjuvant treatment and before surgery, and at 3 and 12 months after surgery were viably frozen and stored in liquid nitrogen in the Duke Substrate Services Core and Research Support facility. PBMCs from 2 healthy donors were similarly utilized for Phytohaemagglutinin-stimulated (PHA-P; Invivogen) and non-stimulated staining controls and viably frozen in aliquots for inclusion with each batch of patient samples to identify any batch effects. PBMC phenotyping was conducted using a 35-marker mass cytometric approach. PBMCs were gently thawed in RPMI with 10% FBS (ThermoFisher) and 50 mg/mL benzonase nuclease (Millipore Sigma). After thawing, PBMCs were treated with Cell-ID Cisplatin in Maxpar PBS (Standard BioTools) for live-dead discrimination. PBMCs were washed and then stained with Fc block (BD Biosciences) in Maxpar Cell Staining Buffer (Standard BioTools) followed by anti-human CD45 with different isotope conjugates utilized as barcodes for the SU2C-SARC032 samples and the spike-in controls. CD45-stained control PBMCs were washed, spiked-in to SU2C-SARC032 samples, and stained with the extracellular antibodies in the panel diluted in Maxpar Cell Staining Buffer (Standard BioTools). PBMC samples were then barcoded with palladium isotopes (Standard BioTools) and pooled prior to fixation and permeabilization with the respective eBioscience 1X solutions (ThermoFisher). Batching was avoided for samples with low post-thaw viability (viability < 70%). Samples were then stained with the intracellular antibodies in the panel diluted in eBioscience 1X permeabilization solution (ThermoFisher). After staining and washing, cells were again fixed with 1.6% formaldehyde and stained overnight with Cell-ID Intercalator in Maxpar Fix and Perm (Standard BioTools) for singlet discrimination. Data acquisition was performed on a Helios mass cytometer (Standard BioTools) according to manufacturer instructions.

Data was debarcoded with the Standard BioTools provided software package and data was analyzed on Cytobank (Beckman). Geometric gating, iridium isotope gating, and cisplatin gating were used to select for viable singlets. Raw CyTOF data was processed using the “flowCore” Bioconductor R package^48^. Marker abundance was subset to the 35 measured markers. Unstimulated and stimulated PBMC spike-in controls were segregated by different CD45 isotope and excluded from clustering. The marker expression was arcsinh transformed via the “CATALYST” package. Clustering was carried out using the Phenograph^49^ based on the top 30 principal components of marker expression. Hierarchical clustering of the median marker expression of each initial cluster resulted in 9 distinct cell populations. This process was repeated exclusively for the identified T cell clusters, which resulted in 11 distinct T cell subtypes. UMAP embeddings were also derived from the principal components of the marker data for visualization. The “compositions“ R package^47^ was used to apply centered log-ratio transformation on the raw CyTOF fractions before downstream analysis.

### STS single-cell RNA sequencing atlas

Previously published UPS, myxofibrosarcoma (MFS), and LPS (including both DDLPS and well differentiated LPS) scRNA-seq samples were downloaded from GEO under accession codes <u>GSE279852</u> and <u>GSE212527</u>, and all analyses were performed in R using the “Seurat” package^50^. Cells expressing hemoglobin genes, which usually represent artifacts or contaminated cells, were removed from further analysis. Tumor and normal cells were annotated for each sample as previously described^19^, then malignant cells, CAFs, myeloid cells, lymphoid cells, and endothelial cells (macro-groups) were identified separately for each histology. Within each histology-specific macro-group, subclusters with an expression profile not consistent with the macro-group of cells analyzed were removed from further analysis.

For each macro-group, the cells from each sample were normalized with SCTransform using the glmGamPoi method^51^ setting vst.flavor to “v2” followed by integration across samples of all histologies using Harmony^52^. Dissociation induced genes (DIGs) obtained from Van Den Brink et al.^53^ and measured using Seurat’s AddModuleScore, confounding from cycling cells using Seurat’s CellCycleScoring, and the percent of mitochondrial genes for each cell were regressed out during SCTransform normalization. Objects were integrated using the top 3000 most variable features excluding DIGs, ribosomal protein genes, T cell receptor genes (for lymphoid cell clustering), *MALAT1*, and cell cycle-specific genes to avoid confounding in dimensionality reduction and clustering. Harmony was run using 15 dimensions and setting the sample of origin as the blocking variable for batch effect correction. UMAP embeddings were obtained using the first 15 Harmony dimensions, and subclusters were obtained by calculating the k-nearest neighbors (k-NN) from a shared nearest-neighbor graph using the Louvain algorithm implemented in the FindClusters function of the Seurat package at different resolutions continuously from 0.4 to 2.0. The “clustree” R package^54^ was used to determine the optimal resolution for each macro-group. To label the final cell states, both FindAllMarkers and limma-voom were used to perform differential gene expression testing and identify marker genes for each subcluster. Seurat’s FindAllMarkers was run with default parameters except for min.pct = 0.25 and only.pos = TRUE. Limma-voom was run after converting the scRNA-seq data to pseudobulks for each sample as previously described^55,56^. Cell states were defined by comparing the identified marker genes within each cluster with the expression profile of known cell states reported in the literature.

### CIBERSORTx deconvolution of bulk RNA-seq data

Separate scRNA-seq signatures for immune and non-immune cells were built as above and used as input to run CIBERSORTx in raw UMI counts format^20^ with pre- and post-treatment bulk RNA-seq tumor samples normalized to counts per million (CPM). CIBERSORTx was then run in single cell mode using S-mode batch correction and setting the number of permutations for p-value calculation to 500. The bulk RNA-seq data was then separately deconvoluted using the TR4 signature matrix, which consists of immune, epithelial, endothelial, and fibroblast cells derived from bulk RNA-seq of flow-sorted cellular populations^20^. The abundance of different cell types recovered using the TR4 signature was used to normalize the abundance of immune and non-immune single-cell states recovered using CIBERSORTx in single-cell mode, so that for each sample, the total cell fractions summed to 1.0. Fractions were then centered log-ratio transformed using the “compositions” R package^47^ before downstream analysis.

### Differential gene expression and gene set enrichment analysis

Differential gene expression analysis was performed using DESeq2^57^ with treatment arm, time point, and the interaction between the treatment arm and time point included in the design. The effect of RT on gene expression (**Fig. 6B**) was analyzed by contrasting gene expression in post-RT vs. pre-treatment samples, and the effect of pembrolizumab on gene expression after treatment (**Fig. 6C**) was analyzed using the interaction term. Gene set enrichment analysis for the Hallmark gene sets was performed for each comparison with the “fgsea” R package with genes ranked by the −log_10_(p-value) multiplied by the sign of the log_2_(fold change).

### T cell receptor repertoire deconvolution

The T cell receptor repertoire for each sample was deconvoluted from raw bulk RNA-seq FASTQ files using TRUST4^27^. A database of sequences for V, J, and C human genes was built from the International ImMunoGeneTics information system and used for analysis^58^.

### Statistical analysis

The primary endpoint of SU2C-SARC032 was disease-free survival. SU2C-SARC032 was designed to detect a 22% increase in 2-year disease free survival from 50% in the control group to 72% in the experimental group with 80% power at a one-sided 0.05 level of significance. The protocol-defined primary statistical analysis plan was to compare disease-free survival between the two treatment groups using a stratified log-rank test in a modified intention-to-treat population that consisted of all evaluable patients. The prespecified significance level for the primary analysis was a one-sided *P* value of 0.05. Secondary outcomes included distant disease-free survival, local recurrence-free survival, overall survival, wound complications, and treatment-related adverse events as assessed by CTCAE v4.0. Exploratory outcomes included characterizing the peripheral and intra-tumoral immune response to combined pembrolizumab and RT and identifying predictors of clinical response to pembrolizumab and RT. On April 15, 2020, the protocol was amended to include a preplanned subgroup analysis to compare primary and secondary endpoints for patients in the control versus experimental arm for each sarcoma immune class. The study was not powered for secondary or exploratory analyses. A run-in safety analysis was performed after the first three patients were accrued to the experimental arm and observed for 30 days after cycle 3 of pembrolizumab to assess for grade 4 or greater toxicities. The primary endpoint analysis was triggered once 45 disease-free survival events were observed, and outcomes were masked to study investigators until this time. The primary and secondary outcomes, including all adverse events were reported previously^12^. Only evaluable patients were included in exploratory analyses, and patients with missing data were excluded.

The DFS interval was defined as the time from randomization to disease recurrence or death. Patients who were alive without documented disease recurrence were censored at the time of their last disease evaluation. In all cases, DFS was calculated using the Kaplan-Meier method, and groups were compared using a two-sided log-rank test. Hazard ratios were obtained by fitting univariable Cox proportional hazards models using the coxph function from the “survival” R package, and the association of each individual variable with DFS was calculated using two-sided Wald tests. Distributions of paired samples were compared using paired Wilcoxon signed-rank tests, and distributions of unpaired samples were compared using two-sided Wilcoxon rank-sum tests. When comparing distributions of more than two groups, Kruskal-Wallis tests were used to test for overall differences among groups, followed by post-hoc Dunn’s tests for pairwise comparisons. Proportions were compared using two-sided Fisher’s exact tests. Correlations between variables were measured using Spearman correlation. Multiple hypothesis testing correction was performed using the Benjamini-Hochberg method. No threshold for significance was prespecified due to the exploratory nature of the correlative analyses, and nominal *P* values were reported. All analyses were performed either in R version 4.3.1 or using Prism 8 (GraphPad Software).

### Data and code availability

The RNA sequencing data generated in this study have been deposited in the Database of Genotypes and Phenotypes at phs003921.v1.p1. The scRNA-seq UPS/LPS atlas was built from previously published data available from GEO through the accession code <u>GSE279852</u> and <u>GSE212527</u>. The code used to perform the analyses reported in the manuscript has been deposited at https://github.com/StefTesta/SARC032_correlative. The EcoTyper code is available for nonprofit academic use at https://github.com/ejmoding/sarcomaecotyper. All other datasets and information necessary to replicate this work are provided in the supplementary tables. Any additional information is available from the corresponding authors upon reasonable request.

## Acknowledgements

We thank the patients and caregivers who participated in this study. We thank Denise Reinke, Steven Young, Scott Okuno, and the research coordinators at SARC for supporting this trial by collecting patient data and samples. In particular, we thank Erin Kozlowski and Meghan Blalock who provided expert project management at SARC. We thank the UNC core for mass cytometry that provided necessary equipment and materials for the CyTOF analysis. We also thank the participating clinicians and clinical research teams at the participating sites for enrolling patients to make this international trial and correlative studies possible. This work was supported by a Stand Up To Cancer Catalyst Research Team grant with support from Merck (DGK, KJW, EJM, and YMM), a research grant from the Investigator-Initiated Studies Program of Merck Sharp & Dohme Corp (DGK), the National Cancer Institute (EJM: K08CA25542501; DGK: R35CA197616), the Department of Defense (EJM: W81XWH-22-1-0161), the My Blue Dots organization (EJM), the Tad and Diane Taube Family Foundation (MvR and EJM), and the GPA Andrew Ursini Charitable Fund (AMH). The opinions expressed in this paper are those of the authors and do not necessarily represent those of Merck Sharp & Dohme Corp. The schematic in Fig.1 was created with BioRender.com.

---

**Fig. S1:**
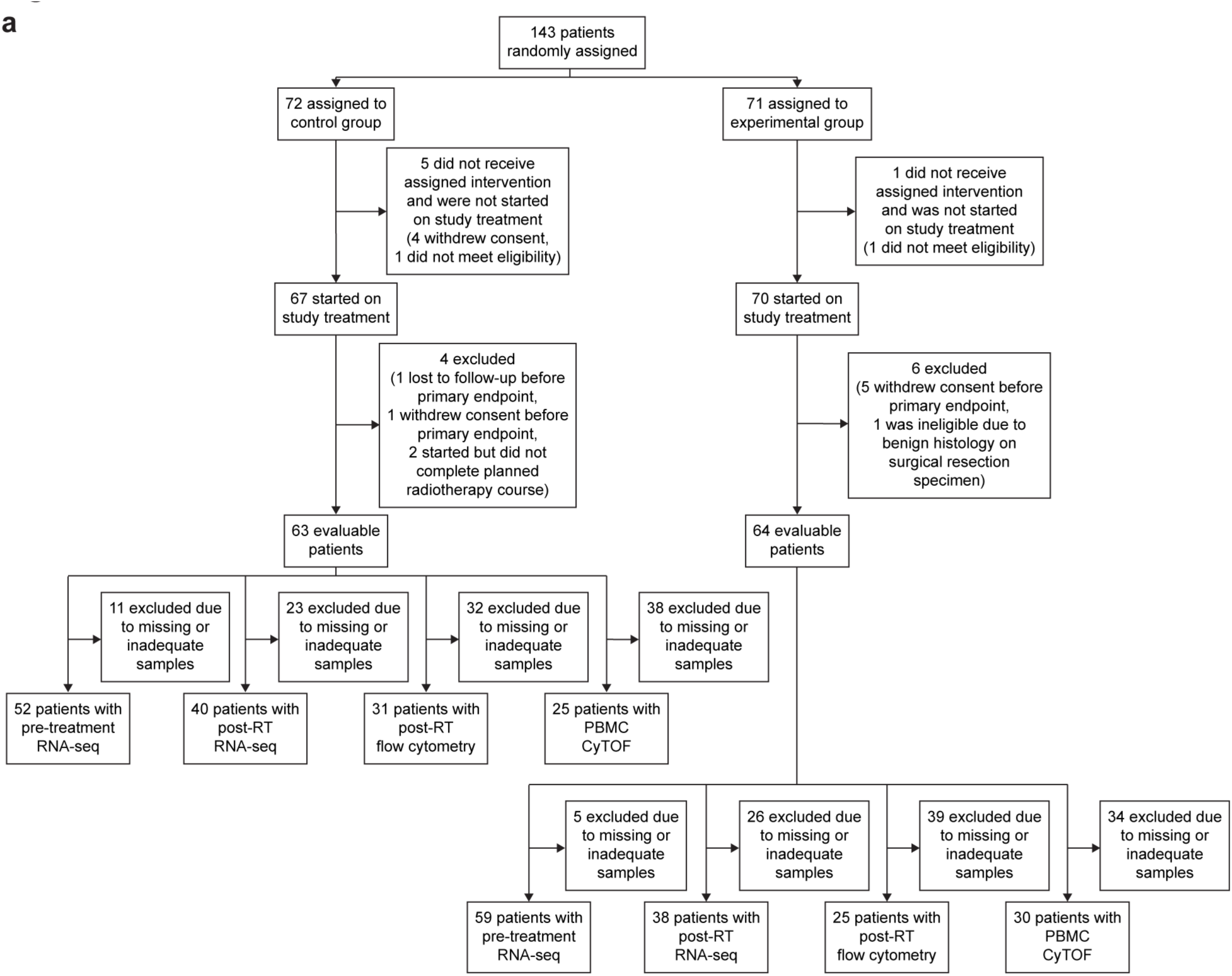
CONSORT diagram. **a,** Flow diagram for the SU2C-SARC032 clinical trial. RT: radiotherapy, RNA-seq: RNA-sequencing, PBMC: peripheral blood mononuclear cells, CyTOF: Cytometry by Time of Flight.

**Fig. S2:**
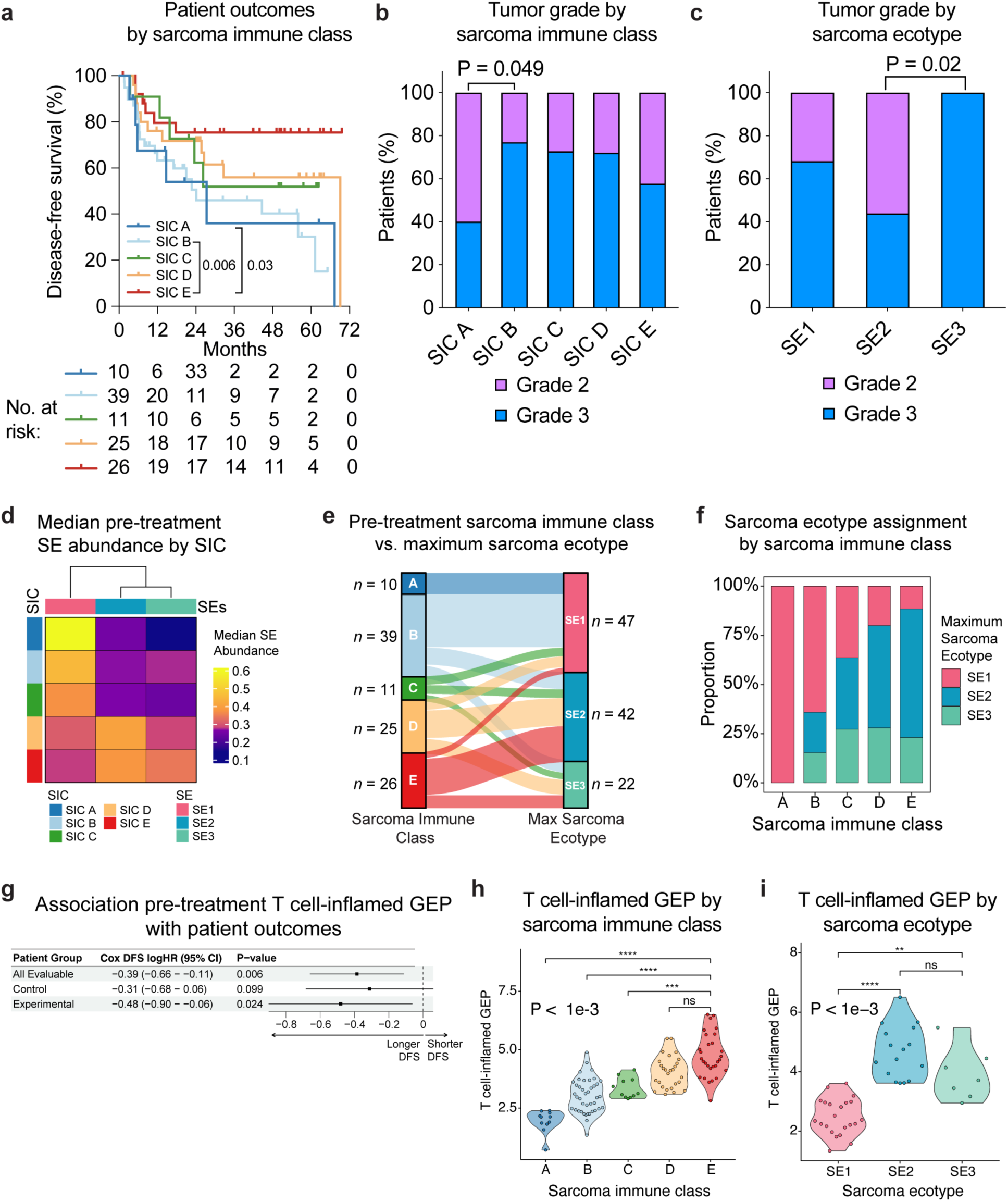
Pre-treatment sarcoma microenvironment gene expression signatures. **a**, Kaplan-Meier curves showing disease-free survival (DFS) by SIC. P-values were calculated using two-sided log-rank tests. **b-c**, Proportion of patients with grade 2 or grade 3 tumors grouped by (**b**) sarcoma immune class (SIC) and (**c**) sarcoma ecotype (SE). P-values were calculated using two-sided Fisher’s exact tests. **d**, Heat map showing median SE abundance in pre-treatment samples for patients classified as SIC A, B, C, D, or E. **e**, River plot showing overlap between SIC and SE classification among pre-treatment samples. Patients are categorized as SE1, SE2, or SE3 based on the maximum SE abundance in the pre-treatment samples. The thickness of the bars represents the proportion of patients in each group. **f**, Bar plot showing proportion of patients assigned to SE1, SE2, or SE3 based on maximum SE abundance in the pre-treatment samples among patients belonging to a different SIC. **g**, Forest plot showing the association of the T cell-inflamed gene expression profile (GEP) described by Ayers et al. (*Journal of Clinical Investigation*, 2017) with DFS by treatment arm calculated using univariable Cox proportional hazards models. P-values were calculated using two-sided Wald tests. **h**-**i**, Violin plots displaying the distribution of T cell-inflamed GEP scores grouped by (**h**) SIC and (**i**) SE. P-values were calculated using Kruskal-Wallis tests and pairwise comparisons between SICs or SEs were computed using two-sided Dunn’s tests (ns: p > 0.05; *: p <= 0.05; **: p <= 1e-2; ***: p<= 1e-3; ****: p <= 1e-4).

**Fig. S3:**
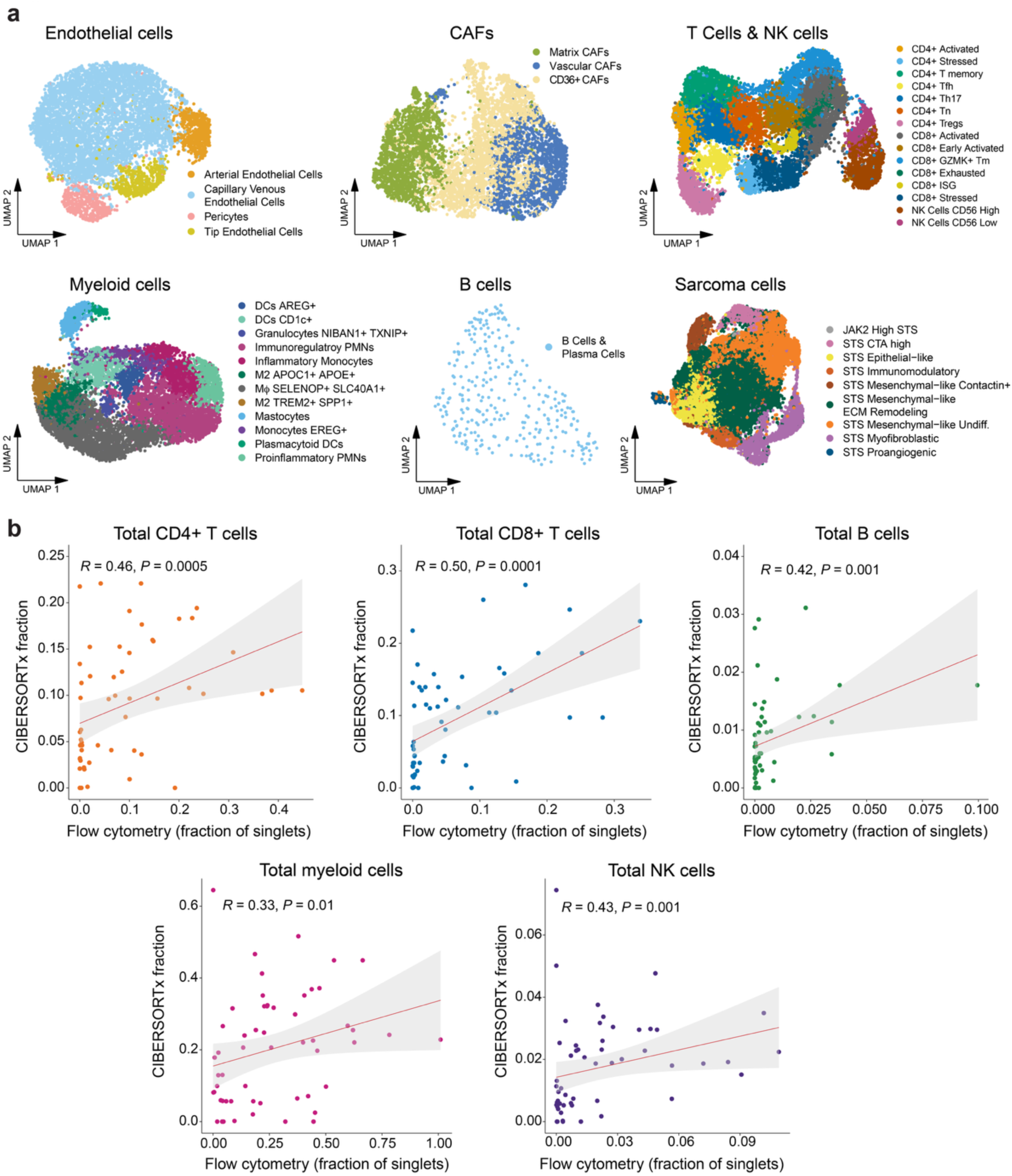
A single cell RNA-seq (scRNA-seq) reference atlas for bulk RNA-seq deconvolution of undifferentiated pleomorphic sarcomas (UPS) and liposarcomas (LPS). **a**, UMAP plots showing the different non-malignant and malignant cell states identified for each major cell type in the STS scRNA-seq reference atlas, including endothelial cells, cancer associated fibroblasts (CAFs), B cells, lymphoid, and myeloid cells. **b**, Scatter plots showing the correlation between the abundance of cell types recovered in bulk RNA-seq of post-treatment formalin-fixed paraffin embedded (FFPE) samples using CIBERSORTx and measured through flow cytometry in the matched dissociated fresh tumor samples. Spearman’s correlation coefficients and two-sided P-values are displayed on the graph. The line of best fit by linear regression and 95% confidence intervals are shown on the graph.

**Fig. S4:**
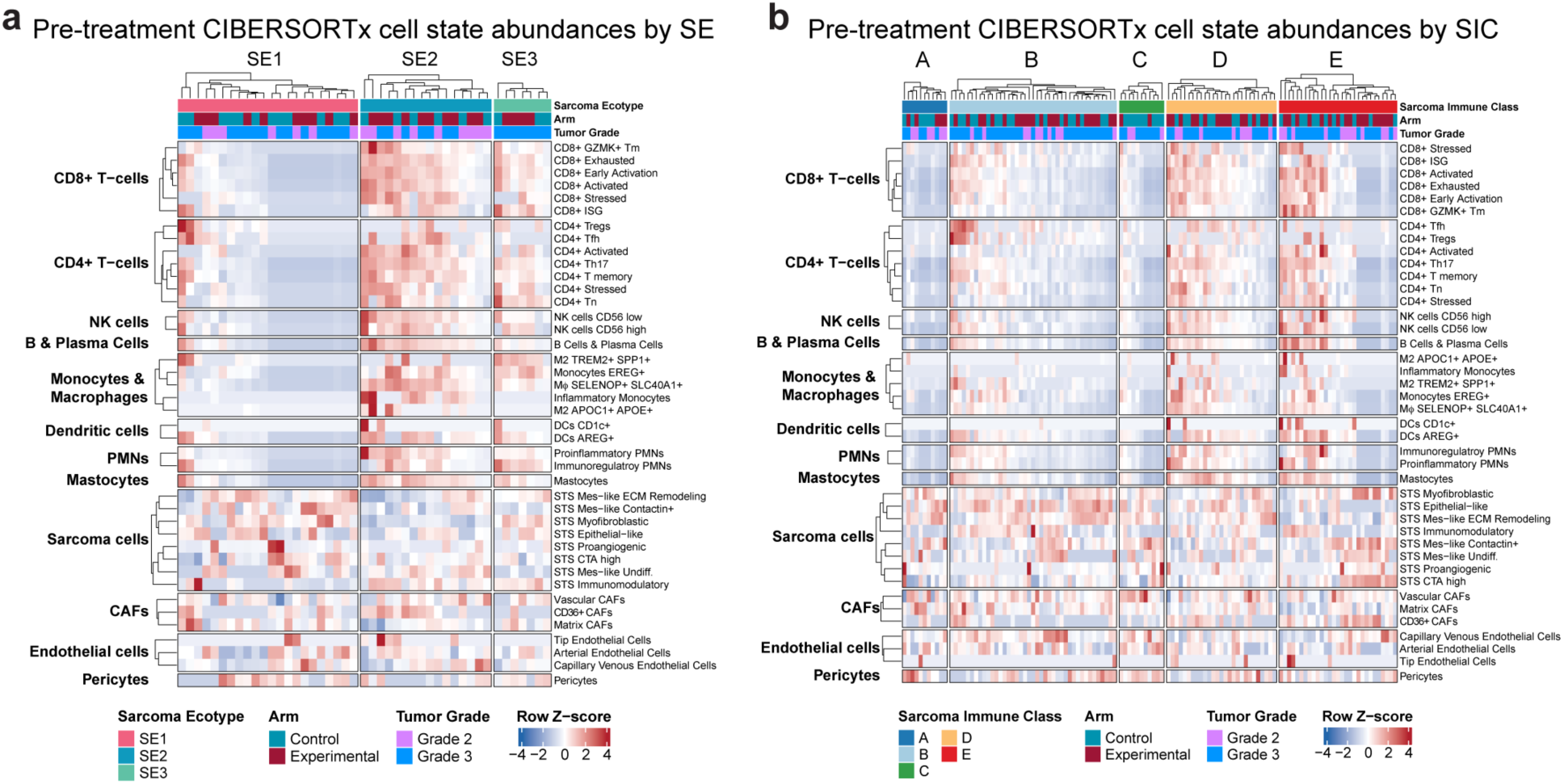
Tumor microenvironment cell states across sarcoma immune classes and sarcoma ecotypes. **a** and **b**, Heat maps showing the abundance of cell states recovered with CIBERSORTx in pre-treatment samples grouped by (**a**) SE and (**b**) SIC. M<Ι: macrophages.

**Fig. S5:**
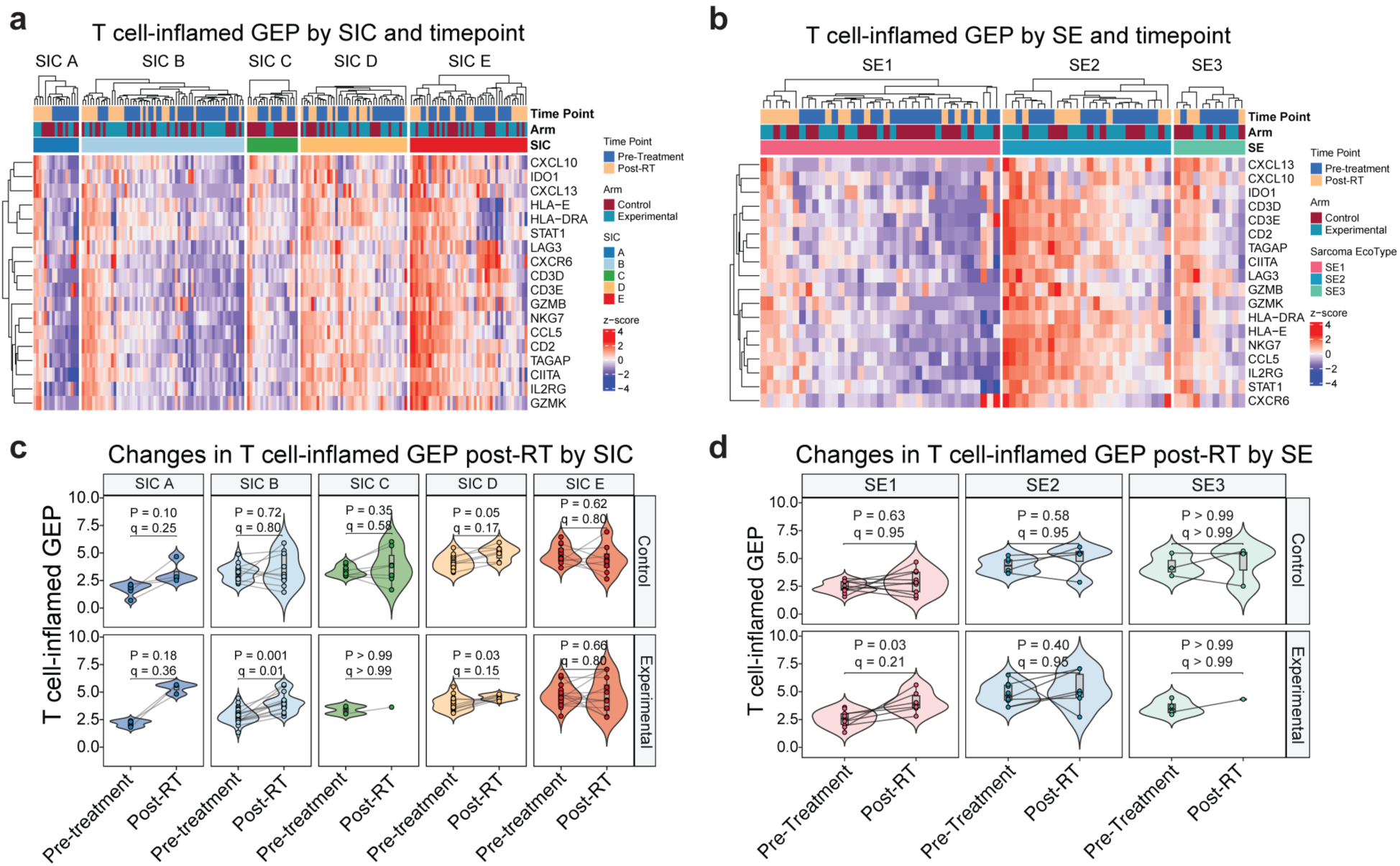
Changes in the T cell-inflamed gene expression profile (GEP) post-radiotherapy (post-RT). **a**-**b**, Heat maps showing the expression of genes in the T cell-inflamed GEP score described by Ayers et al. (*Journal of Clinical Investigation*, 2017) in pre-treatment and post-RT samples grouped by pre-treatment (**a**) sarcoma immune class (SIC) and (**b**) sarcoma ecotype (SE). **c-d**, Box-violin plots displaying the change in T cell-inflamed GEP score from pre-treatment to post-RT grouped by (**c**) sarcoma immune class and treatment arm and (**d**) sarcoma ecotype and treatment arm. P-values were calculated using two-sided Wilcoxon signed-rank tests and adjusted for multiple hypothesis testing using the Benjamini-Hochberg method.

**Fig. S6:**
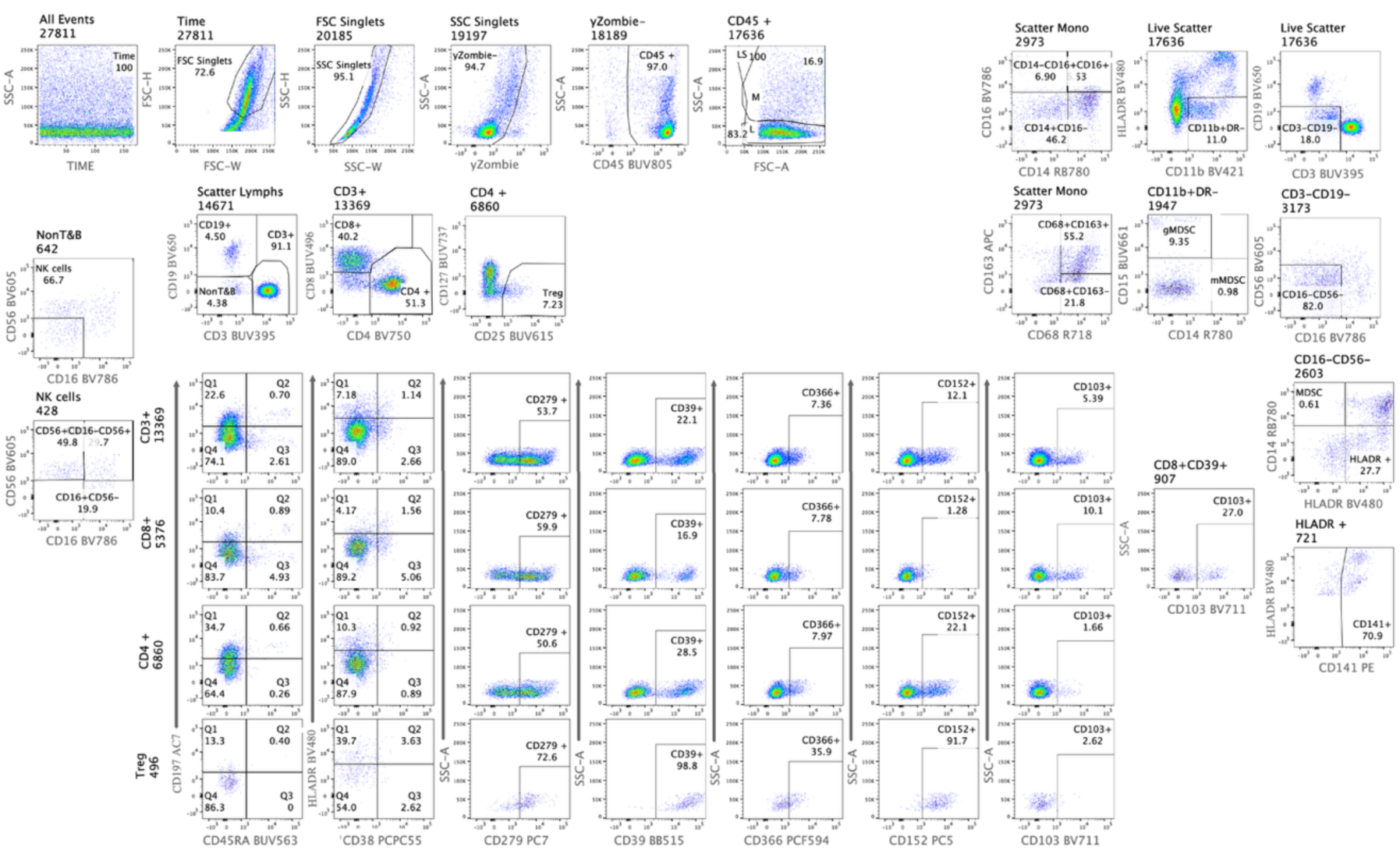
Flow cytometry gating strategy for post-treatment tumor samples.

**Fig. S7:**
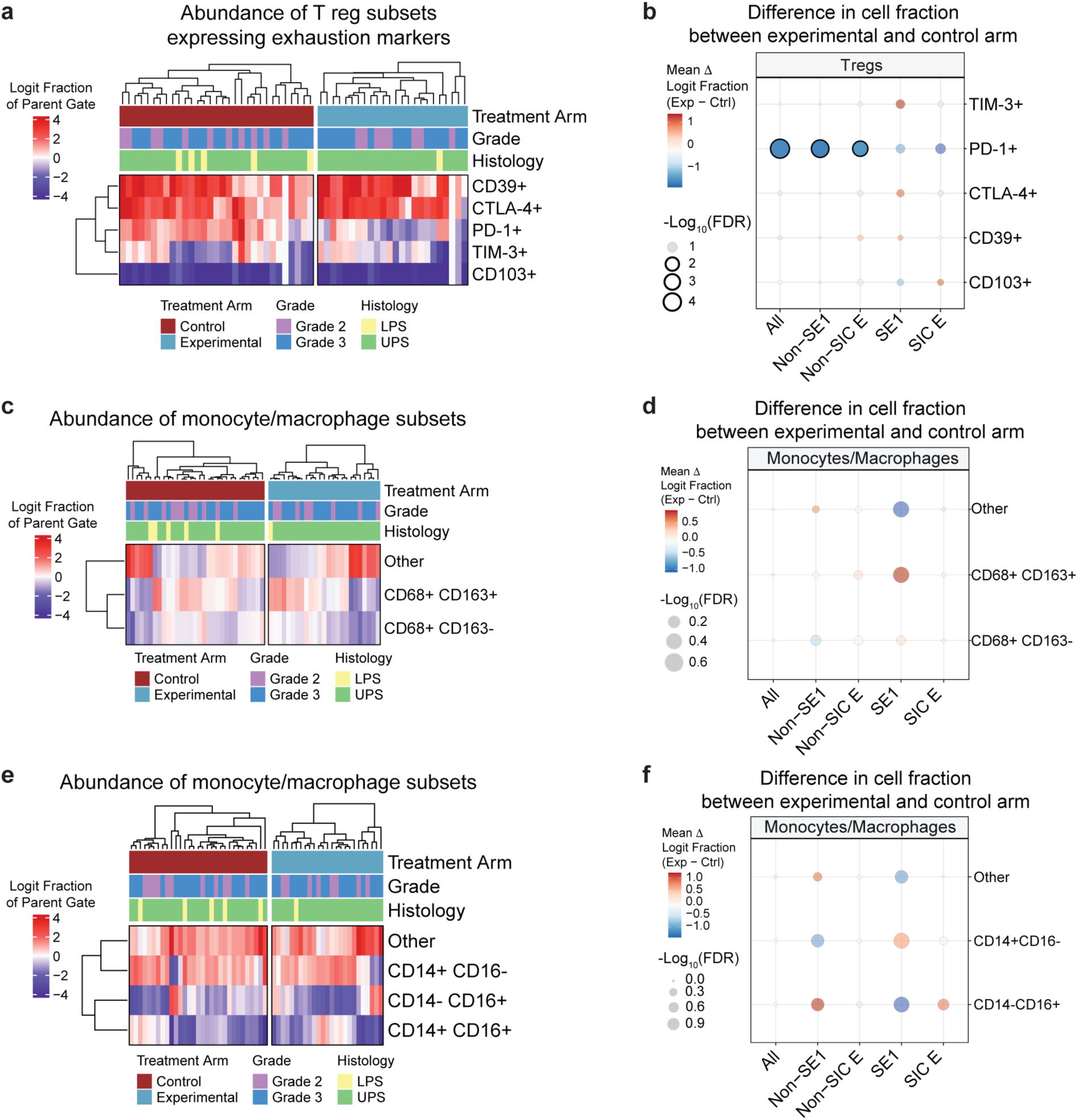
Flow cytometry of post-treatment samples. **a**, Heat map showing the abundance of T regulatory cells (T reg) subsets expressing different exhaustion markers in resected sarcomas after radiotherapy with or without pembrolizumab measured by flow cytometry. Patient samples are grouped according to study arm. Cell type abundances are reported as logit normalized fractions of the total number of cells in the parent gate. **b**, Dot plot showing mean difference in abundance of different T regs subsets, measured as logit fraction of parent gate, between patients in the experimental and control arm. Samples are grouped based on baseline SE1 and SIC E classification. P-values were calculated using two-sided Wilcoxon rank-sum tests. Multiple hypothesis testing correction was applied using the Benjamini-Hochberg method. **c**, Heat map showing the abundance of monocytes/macrophages subsets expressing CD68 and/or CD163 in resected sarcomas after radiotherapy with or without pembrolizumab measured by flow cytometry. Patient samples are grouped according to study arm. Cell type abundances are reported as logit normalized fractions of the total number of cells in the parent gate. **d**, Dot plot showing mean difference in abundance of monocytes/macrophages subsets expressing CD68 and/or CD163, measured as logit fraction of parent gate, between patients in the experimental and control arm. Samples are grouped based on baseline SE1 and SIC E classification. P-values were calculated using two-sided Wilcoxon rank-sum tests. Multiple hypothesis testing correction was applied using the Benjamini-Hochberg method. **e**, Heat map showing the abundance of monocytes/macrophages subsets expressing CD14 and/or CD16 in resected sarcomas after radiotherapy with or without pembrolizumab measured by flow cytometry. Patient samples are grouped according to study arm. Cell type abundances are reported as logit normalized fractions of the total number of cells in the parent gate. **f**, Dot plot showing mean difference in CLR abundance of monocytes/macrophages subsets expressing CD14 and/or CD16, measured as logit fraction of parent gate, between patients in the experimental and control arm. Samples are grouped based on baseline SE1 and SIC E classification. P-values were calculated using two-sided Wilcoxon rank-sum tests. Multiple hypothesis testing correction was applied using the Benjamini-Hochberg method.

